# Comprehensive genetic and functional analysis of FcγRs in rituximab therapy for autoimmunity reveals a key role for FcγRIIIa on NK cells

**DOI:** 10.1101/2021.08.25.21262612

**Authors:** James I Robinson, Md Yuzaiful Md Yusof, Vinny Davies, Dawn Wild, Michael Morgan, John C Taylor, Yasser El-Sherbiny, David L Morris, Lu Liu, Andy C Rawstron, Maya H Buch, Darren Plant, Heather J Cordell, John D Isaacs, Ian N Bruce, Paul Emery, Anne Barton, Timothy J Vyse, Jennifer H Barrett, Edward M Vital, Ann W Morgan, on behalf of the MATURA, MASTERPLANS Consortia

**Author notes:** Please see Appendix I (on behalf of the MATURA). Please see Appendix II. These authors contributed equally to this work and should be considered to have equal authorship status (James I Robinson, Md Yuzaiful Md Yusof). These authors should be considered as having joint senior authorship status (Timothy J Vyse, Jennifer H Barrett, Edward M Vital, Ann W Morgan). Corresponding Author Professor Ann W Morgan Leeds Institute of Cardiovascular and Metabolic Medicine School of Medicine, LIGHT Building, Clarendon Way University of Leeds, Leeds LS2 9DA, UK.

## Abstract

B cell depletion using rituximab is widely used to treat autoimmune diseases, but patient response varies. The efficacy of rituximab is limited by the efficiency of depletion. Strategies to improve response include altering rituximab dosing, switching anti-CD20-mAb, alternative B cell targets, or non-B cell targeted therapies. Implementing an appropriate strategy requires understanding of the mechanism(s) of resistance to depletion and, if this varies between individuals, a means to test for it. Rituximab kills B cells via a variety of Fcγ receptor (FcγR)-dependent mechanisms, including antibody-dependent cellular cytotoxicity (ADCC), as well as non-FcγR mechanisms. We conducted a longitudinal cohort study in rheumatoid arthritis (RA) and systemic lupus erythematosus (SLE) using two national registries. Qualitative and quantitative *FCGR* functional variants were measured using multiplexed ligation-dependent probe amplification, supplemented by novel *FCGR2C* assays.

We provide consistent evidence that *FCGR3A,* specifically increased number of copies of the *FCGR3A-*158V allele, was the major FcγR gene associated with rituximab response, including clinical response in RA and SLE and depth of B cell depletion in the combined cohort. In SLE, we provide preliminary data suggesting increased *FCGR2C* ORF copies were also associated with improved clinical response. Furthermore, we demonstrated the impact of disease status and concomitant therapies on both natural killer cell FcγRIIIa expression and rituximab-induced ADCC; demonstrating increased FcγRIIIa expression and *FCGR3A* genotype were independently associated with clinical response and B cell depletion. Our findings highlight the importance of enhancing FcγR-effector functions, may help stratify patients, and support ongoing development of next-generation CD20 depleting therapeutics.

**One Sentence Summary:** The high affinity FcγRIIIa allotype on NK cells explains depth of B cell depletion and clinical response in rituximab therapy for autoimmune disease

## INTRODUCTION

Rheumatoid arthritis (RA) and systemic lupus erythematosus (SLE) are two autoimmune diseases where B cells are considered to play a central role in disease pathogenesis through autoantibody production, antigen presentation and pro-inflammatory cytokine expression (*1, 2*). Depletion of naive, memory and malignant B cells using CD20 monoclonal antibodies (mAb) is a therapeutic strategy that has been exploited for the treatment of haematological malignancies and autoimmune diseases. Rituximab, the first molecule in this class, is the top-selling biologic in oncology and second highest across all medical indications, with use continuing to expand due to new indications and the recent licensing of cheaper biosimilars (*3*). In RA, rituximab is licensed for patients with active inflammation despite therapy with disease modifying antirheumatic drugs (DMARDs) (*4*). In SLE, rituximab therapy is recommended by the European League Against Rheumatism (EULAR) for use in severe and refractory disease (*5–9*). Despite the success of this therapeutic strategy, clinical responses are variable and there remains an unmet need to understand the mechanisms of sub-optimal response to improve outcomes at an individual patient level. Only by meeting this need can rational strategies to improve clinical outcomes be proposed. A substantial body of evidence indicates an association between achieving complete peripheral B cell depletion and clinical response in RA and SLE, particularly when highly sensitive flow cytometry (HSFC) assays are used to enumerate circulating B cells (*10, 11*). However, mechanistic studies to explain this variability in depth of depletion in autoimmunity are limited (*12*).

Rituximab is a chimeric anti-CD20 mAb, with a native IgG1 Fc, which crosslinks Fcγ receptors (FcγRs) expressed on immune effector cells. Potential explanations for variability in depletion invoke three main mechanisms of B cell killing, and the relative importance of each may differ between autoimmune and malignant diseases. These include antibody dependent cellular cytotoxicity (ADCC), complement-dependent cytotoxicity and direct signalling-induced cell death, with variable evidence available from animal models, *in vitro* and clinical studies (*13*). Genetic studies in human autoimmune and malignant diseases, outlined in more detail below, provide strong evidence for natural killer (NK) cell-mediated ADCC, delivered through FcγRIIIa as the principal mechanism of B cell depletion.

The expression of FcγR subtypes is known to differ between leukocyte populations. NK cells are generally characterised by expression of FcγRIIIa, with greater expression in the circulation than in tissues. In the latter, other FcγR-mediated mechanisms and innate cells may also contribute, such as antibody-dependent cellular phagocytosis, which leads to clearance of rituximab-opsonised B cells by cells of the reticuloendothelial system, tissue macrophages or neutrophils (*14*). NK cells from some individuals express FcγRIIc, or rarely FcγRIIb dependent upon specific gene rearrangements (*15*). Phagocytic cells express activatory FcγRs (i.e. FcγRIIa, FcγRIIc, FcγRIIIa and FcγRIIIb) in a cell-type specific manner. There is a single inhibitory FcγR (FcγRIIb), which fine-tunes activatory signals. FcγRIIb is the only FcγR expressed on B cells where it may contribute to rituximab-mediated CD20 internalisation. Innate cell activation may also lead to release of soluble mediators that modulate FcγR expression on phagocytic cells, for example interferon gamma (IFNγ) release from activated NK cells and complement component 5a (C5a) release from Kupffer cells exposed to IgG-coated B cells, potentially further enhancing FcγR-mediated clearance mechanisms (*16–18*). Differences in the relative importance of these mechanisms between different disease states may be explained by inherent differences in IgG and immune complex structure, B cell biology, immune effector functions and concomitant immunosuppressive medications.

Recent evolutionary gene duplications and rearrangements have created a structurally variable genetic locus encoding the FcγRs, with both gene duplications and deletions observed (*15, 19, 20*). Consequently, due to the high homology between paralogs, genotyping of the low-affinity *FCGR* locus is technically challenging. There are well-described functional variants that affect FcγR-IgG interaction affinity and/or FcγR expression, which have the potential to modulate a multitude of IgG effector functions that ultimately lead to rituximab-induced B cell depletion and downstream clinical response. Some published studies have evaluated genetic predictors of response to rituximab in genes encoding the low affinity FcγRs in haematological malignancies (*21–23*), RA (*24–28*), SLE (*29*) and treatment-related complications, such as rituximab-induced neutropenia (*30, 31*). Candidate variants evaluated included *FCGR3A* (F158V, rs3969910) and *FCGR2A* (H131R, rs1801274), which encode receptors with a single amino acid difference in the IgG binding site. The FcγRIIIa-158V and FcγRIIa-131H allotypes have increased affinity for IgG1 and IgG2, respectively. However, in general, these genetic studies have lacked statistical power (sample sizes ranging from 12 to 212 patients), included heterogeneous patient cohorts, treatment regimens, and clinical outcome measures, as well as genotyping technologies that were neither comprehensive nor accounted for copy number variation (CNV), which can confound meta-analyses (*25–28*). We and others have shown that copy number of *FCGR3A* (*32*) and *FCGR3B* (*33, 34*) correlate with cell surface expression, which may further modulate ADCC and B cell depletion.

To provide a mechanistic explanation for the clinical importance of rituximab-FcγR engagement on clinical response, and the intermediate phenotype of depth of B cell depletion, we performed a comprehensive analysis of functional variants within the low affinity *FCGR* genetic locus that addresses the complexity at this locus. We evaluated qualitative variation that modulates rituximab-FcγR binding affinity and quantitative CNV influencing FcγR cell surface expression. In order to make replication of our work simpler, we have adopted a commercial multiplexed ligation-dependent probe amplification (MLPA) platform for measuring genetic variation at the *FCGR* locus, supplemented with our in-house assays, which together offer combined assays for qualitative single nucleotide polymorphisms (SNPs) and quantitative CNVs (*32*). The conventional approach to analysing *FCGR3A* F158V genotyping data has been to treat it as a simple two copy biallelic SNP, whereas our genotyping approach provided the opportunity to take account of CNV and explore both gene and allele copy number in RA and SLE. Moreover, in combination with our detailed analysis of clinical predictors of response in large patient cohorts, our results will help to facilitate the ultimate goal of personalizing rituximab therapy in autoimmune diseases.

## RESULTS

### Baseline clinical characteristics and association with clinical response to rituximab and complete B cell depletion in RA

Prior to genetic and functional FcγR studies, we analysed clinical, B cell, and routine clinical immunological associations with B cell depletion and clinical response to determine key covariates for subsequent analyses.

The baseline characteristics of our RA cohort were typical of patients receiving biologics in the UK (Table 1). The mean age at first RTX treatment was 59 years, with disease duration 12 years; 78% were female, 97% of European descent and 91.5% were either rheumatoid factor (RF) and/or anti-citrullinated peptide antibody (ACPA) positive. The majority for whom data were available in electronic patient records (74%) received concomitant DMARDs (hydroxychloroquine, methotrexate, leflunomide), and had previously received a TNFi (71%).

**Table 1.**
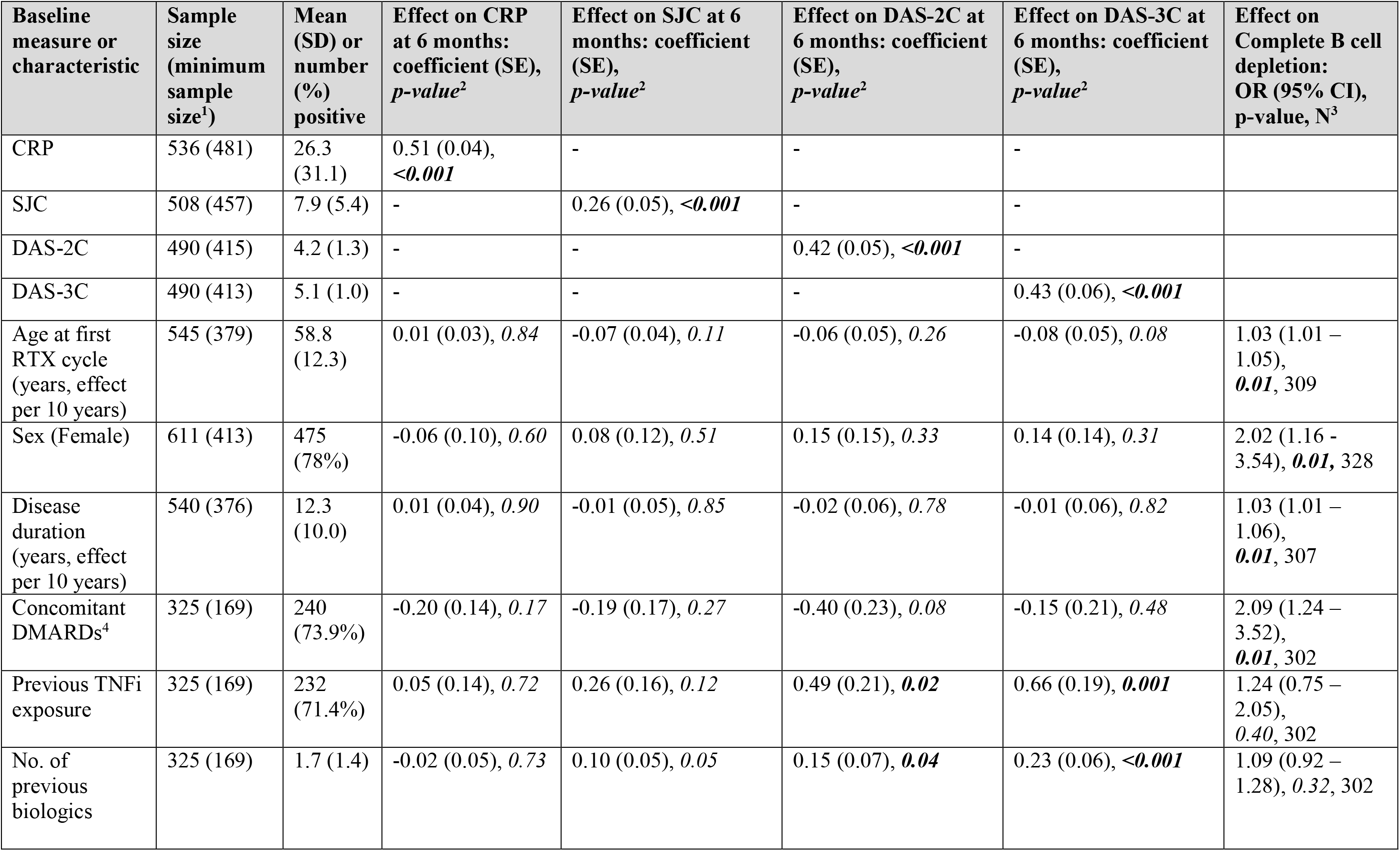
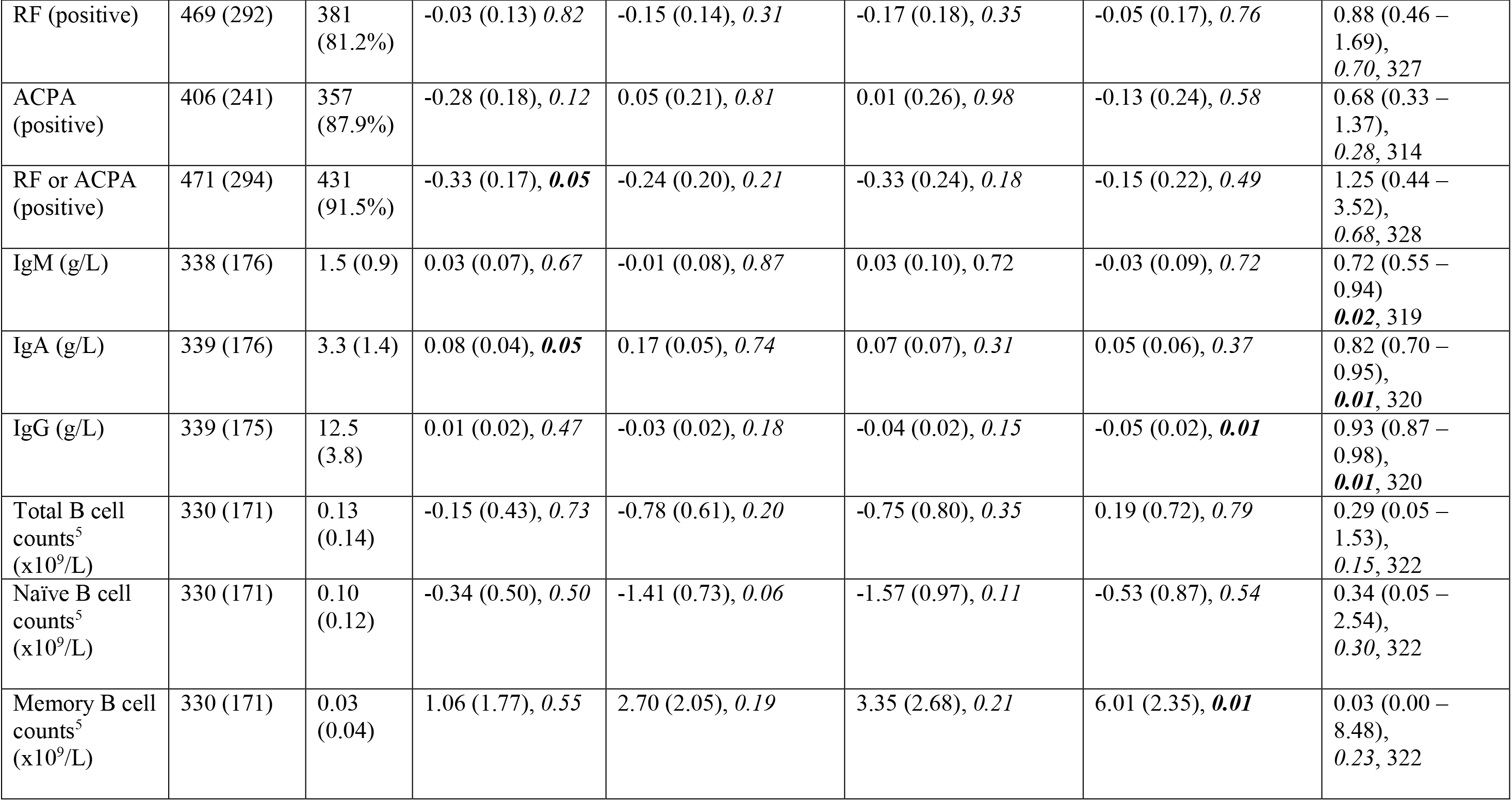
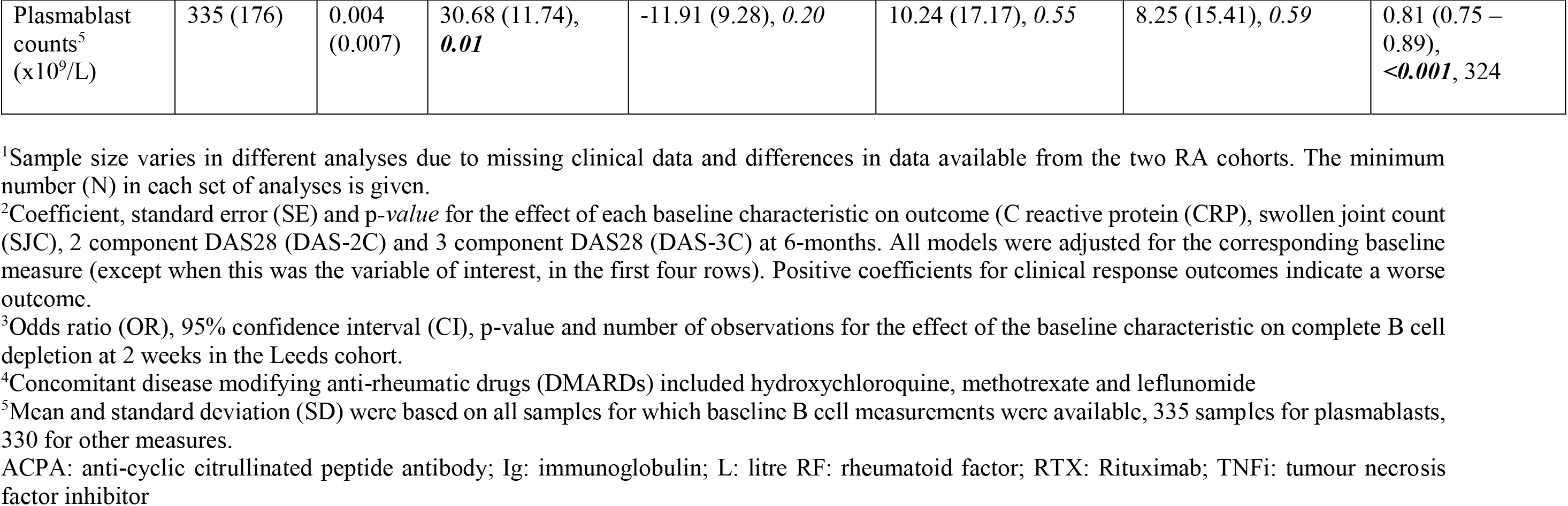
Baseline clinical characteristics, laboratory measures and association with clinical outcomes and complete B cell depletion in rheumatoid arthritis.

As expected, 6-month measures of clinical response to rituximab (i.e. C-reactive protein (CRP), swollen joint count in 28 joints (SJC28), 2-component disease activity score in 28 joints using CRP (2C-DAS28CRP) and 3-component disease activity score in 28 joints using CRP (3C-DAS28CRP)) were highly associated with the corresponding baseline measures (p<0.001). Higher baseline IgG was associated with lower 3C-DAS28CRP (p=0.01) at 6 months, while previous exposure to TNFi (p=0.001), number of previous biologics (p<0.001) and higher baseline memory B cell count (p=0.01) were associated with higher 3C-DAS28CRP at 6 months. No clear associations were observed between other salient baseline clinical and serological markers and clinical response at 6 months, after adjusting for the corresponding baseline measures Table 1).

Circulating B cell counts were analysed using HSFC at 0 and 2 weeks and recorded in 330 patients from the RA cohort treated in Leeds. Complete depletion 2 weeks after rituximab therapy was achieved in 192/328 (58.5%) patients. Most of the remaining patients had low, but detectable, post-rituximab total B cell counts, median (IQR) 0.0009 (0.0005 to 0.0021) x 10^9^/L. Baseline factors associated with increased odds of achieving complete depletion were older age at first rituximab infusion (p=0.01), female gender (p=0.02), longer RA disease duration (p=0.01), and concomitant use of DMARDs (p=0.01). Higher immunoglobulin (IgM, IgA and IgG) levels (p=0.02, 0.01, 0.01, respectively) and higher plasmablast counts at baseline (p<0.001) were associated with lower odds of complete depletion Table 1).

### Baseline clinical characteristics and association with clinical response to rituximab and complete B cell depletion in SLE

The baseline characteristics of the SLE cohort (n=262) are described in Table 2. Mean age was 40 years at first rituximab infusion, 91% were female, 61.5% were Caucasian, 52.5% were anti-double stranded deoxyribonucleic acid (dsDNA) antibody positive, 69.9% were anti-extract nuclear antigen (ENA) antibody positive, and the most common manifestations requiring rituximab therapy were mucocutaneous (49%), musculoskeletal (46%) and renal (43%).

**Table 2.** Baseline clinical characteristics, laboratory measures and association with clinical outcomes and depletion in systemic lupus erythematosus.

| Baseline measure or characteristic | Sample Size<sup>1</sup> | Mean (SD) or number (%) positive | Effect on BILAG response (Major or Partial Clinical Response) at 6 months: OR (95% CI), *p-value*, N<sup>2</sup> | Effect on BILAG Major Clinical Response at 6 months: OR (95% CI), *p-value*, N<sup>2</sup> | Effect on complete B cell depletion: OR (95% CI), *p-value*, N<sup>2</sup> |
| --- | --- | --- | --- | --- | --- |
| Age at first RTX cycle (years effect per 10 Years) | 262 | 40 (14) | 0.81 (0.67 – 0.98), **0.03**; 262 | 0.88 (0.73 – 1.06), *0.17*, 262 | 0.91 (0.67 – 1.24), *0.56*, 85 |
| Sex (Female) | 262 | 238 (91%) | 1.55 (0.66 – 3.66), *0.31*, 262 | 1.05 (0.43 – 2.56), *0.91*, 262 | 0.53 (0.05 – 6.02), *0.61*, 85 |
| Ethnicity | 262 |  |  |  |  |
| Caucasian |  | 161 (61.5%) | 1.06 (0.62 – 1.80), *0.84*, 262<sup>3</sup> | 1.18 (0.70 – 1.98), *0.54*, 262<sup>3</sup> | 1.13 (0.45 – 2.84), *0.80*, 85<sup>3</sup> |
| South Asian |  | 39 (14.9%) |  |  |  |
| Chinese/SE Asian |  | 13 (5.0%) |  |  |  |
| Afro-Caribbean |  | 31 (11.8%) |  |  |  |
| Mixed/Undisclosed |  | 18 (6.8%) |  |  |  |
| Disease Duration at first RTX cycle (years, effect per year) | 261 | 8 (6) | 0.99 (0.96 – 1.02), *0.56*, 261 | 0.99 (0.96 – 1.03), *0.64*, 261 | 1.03 (0.96 – 1.10), *0.45*, 85 |
| Concomitant DMARDs | 85 | 60 (70.6%) | 0.69 (0.22 – 2.14), *0.52*, 85 | 0.81 (0.31 – 2.11), *0.66*, 85 | 0.99 (0.39 – 2.51), *0.98*, 85 |
| Concomitant anti-malarials, | 262 | 225 (85.9%) | 1.32 (0.64 – 2.72), *0.45*, 262 | 0.96 (0.46 – 1.99), *0.91*, 262 | 1.94 (0.67 – 5.61), *0.22*, 85 |
| Concomitant oral prednisolone | 262 | 193 (73.7%) | 0.80 (0.44 – 1.46), *0.48*, 262 | 1.06 (0.59 – 1.90), *0.84*, 262 | 1.10 (0.42 – 2.90), *0.85*, 85 |

| Baseline measure or characteristic | Sample Size<sup>1</sup> | Mean (SD) or number (%) positive | Effect on BILAG response (Major or Partial Clinical Response) at 6 months: OR (95% CI), *p*-value, N<sup>2</sup> | Effect on BILAG Major Clinical Response at 6 months: OR (95% CI), *p*-value, N<sup>2</sup> | Effect on complete B cell depletion: OR (95% CI), *p*-value, N<sup>2</sup> |
| --- | --- | --- | --- | --- | --- |
| **Total No. of positive autoantibodies** | 186 | 1.9 (1.3) | 1.13 (0.89 – 1.43), 0.33, 186 | 1.02 (0.82 – 1.28), 0.86, 186 | 0.80 (0.57 – 1.12), 0.19, 85 |
| **anti-Ro** | 199 | 98 (49.2%) | - | - | - |
| **anti-La** | 199 | 38 (19.1%) |  |  |  |
| **anti-Sm** | 194 | 55 (28.4%) |  |  |  |
| **anti-RNP** | 198 | 67 (33.8%) |  |  |  |
| **Anti-dsDNA positive** | 261 | 137 (52.5%) | 1.33 (0.79 – 2.25), 0.28, 261 | 1.13 (0.68 – 1.88), 0.65, 261 | 0.71 (0.30 – 1.67), 0.43, 85 |
| **ENA positive** | 186 | 130 (69.9%) | 1.02 (0.51 – 2.01), 0.96, 261 | 1.05 (0.55 – 2.02), 0.88, 261 | 0.54 (0.21 – 1.36), 0.19, 81 |
| **Low C3 and/or C4 titre** | 261 | 120 (46%) | 1.49 (0.88 – 2.53), 0.14, 261 | 1.57 (0.94 – 2.63), 0.08, 261 | 0.35 (0.14 – 0.88), 0.03, 85 |
| **Immunoglobulin (g/L)** | 238 |  |  |  |  |
| **IgM** |  | 1.33 (1.9) | 0.90 (0.76 – 1.07), 0.22, 238 | 0.99 (0.86 – 1.15), 0.93, 238 | 1.52 (0.82 – 2.82), 0.18, 78 |
| **IgA** |  | 3.88 (1.9) | 0.94 (0.78 – 1.12), 0.49, 238 | 0.98 (0.84 – 1.14), 0.76, 238 | 0.70 (0.47 – 1.06), 0.09, 78 |
| **IgG** |  | 16.9 (5.4) | 0.97 (0.93 – 1.01), 0.12, 238 | 1.00 (0.98 – 1.01), 0.60, 238 | 0.95 (0.88 – 1.02), 0.13, 78 |
| **ESR (mm/h)** | 192 | 30.4 (27) | 0.99 (0.98 – 1.00), 0.05, 192 | 1.00 (0.99 – 1.01), 0.88, 192 | 0.99 (0.97 – 1.01), 0.30, 62 |
| **Total B cell counts (x 10<sup>9</sup>/L)** | 73 | 0.1263 (0.13) | 1.00 (1.00 – 1.01), 0.74, 73 | 1.00 (1.00 – 1.01), 0.09, 73 | 1.00 (1.00 – 1.00), 0.79, 73 |
| **Naïve B cell counts (x 10<sup>9</sup>/L)** | 71 | 0.0928 (0.09) | 1.00 (0.99 – 1.01), 0.99, 71 | 1.00 (1.00 – 1.01), 0.17, 71 | 1.00 (0.99 – 1.00), 0.54, 71 |
| **Memory B cell counts (x 10<sup>9</sup>/L)** | 71 | 0.0292 (0.07) | 1.00 (0.99 – 1.02), 0.68, 71 | 1.02 (0.99 – 1.04), 0.22, 71 | 1.00 (0.99 – 1.01), 0.71, 71 |
| Plasmablast counts (x 10<sup>9</sup>/L) | 71 | 0.0054 (0.01) | 0.98 (0.90 – 1.06), 0.60, 71 | 0.95 (0.87 – 1.04), 0.23, 71 | 0.88 (0.80 – 0.98), 0.02, 71 |
| Global BILAG score | 262 | 22 (9.7) | 1.00 (0.97 – 1.02), 0.83, 262 | 1.00 (0.97 – 1.03), 0.93, 262 | 1.00 (0.96 – 1.04), 0.96, 85 |
| SLEDAI-2K score | 262 | 10.8 (5.7) | 1.06 (1.00 – 1.11), 0.03, 262 | 1.02 (0.98 – 1.07), 0.40, 262 | 0.97 (0.90 – 1.05), 0.46, 85 |
| Active BILAG domains (either A or B Grade) | 262 |  |  |  |  |
| General |  | 36 (14.1%) | - | - | - |
| Mucocutaneous |  | 129 (49.2%) |  |  |  |
| Neurology |  | 52 (19.8%) |  |  |  |
| Musculoskeletal |  | 121 (46.2%) |  |  |  |
| Cardiorespiratory |  | 44 (17.2%) |  |  |  |
| Gastrointestinal |  | 18 (6.9%) |  |  |  |
| Ophthalmic |  | 9 (3.4%) |  |  |  |
| Renal |  | 114 (43.1%) |  |  |  |
| Haematology |  | 23 (9.5%) |  |  |  |

<sup>1</sup>Sample size varies in different analyses due to missing clinical data. The minimum number (N) in each set of analyses is given.
<sup>2</sup>Odds ratio (OR), 95% confidence interval (CI), *p*-value and number of observations for the effect of the baseline characteristic on clinical response measures or complete B cell depletion at 2 weeks. Peripheral blood B cell depletion data were only available for the Leeds cohort.
<sup>3</sup>Due to sample size for each ethnicity category, comparison was made between Caucasian vs Non-Caucasian and Caucasian was the reference.
BILAG: British Isles Lupus Assessment Group; dsDNA: double stranded deoxyribonucleic acid; DMARD: disease-modifying anti-rheumatic drug; ENA: extract nuclear antigen; ESR: erythrocyte sedimentation rate; RNP: ribonucleoprotein; RTX: rituximab; SD: standard deviation; SE: South East; SLEDAI-2K: Systemic Lupus Erythematosus Disease Activity Index version 2000

The 6-month measures of clinical response to rituximab in SLE were the British Isles Lupus Assessment Group (BILAG) response (major clinical response (MCR) or partial clinical response (PCR)) and BILAG MCR. At 6 months, 177/262 (67.6%) of patients achieved a BILAG response and 90/262 (34.4%) achieved a BILAG MCR following their first rituximab cycle. Higher SLE Disease Activity Index version 2000 (SLEDAI-2K) score was associated with increased odds of BILAG response at 6 months (p=0.03), while older age reduced the odds of BILAG response at 6 months (p=0.03). No other baseline clinical or serological markers were associated with clinical response Table 2).

Peripheral B cells were analysed using HSFC at 0 and 6 weeks and available in 85 SLE patients who were treated in Leeds. At 6 weeks, 44/85 (51.8%) achieved complete B cell depletion. Low complement (C3 and/or C4) (p=0.03) and higher plasmablast counts at baseline (p=0.02) were associated with reduced odds of complete B cell depletion Table 2).

### Novel FCGR2C QSV assay and functional interpretation of FCGR2C genotyping

To supplement the MLPA panels, we developed a novel *FCGR2C* QSV assay to more accurately determine *FCGR2C* copy number. To biologically validate this assay, we sought to determine whether expression of FcγRIIc (CD32) on NK cells was associated with the number of copies of classical *FCGR2C* ORF alleles (*15*). As a control comparison, we determined NK cell CD32 expression in individuals with a specific *FCGR* locus rearrangement that has previously been shown to encode FcγRIIb expression on NK cells (*20*). This CNR1 deletion (Figure 1A) involves a deletion of both *FCGR2C* and *FCGR3B* with the remaining *FCGR2C* gene bearing a STP allele.

**Figure 1.**
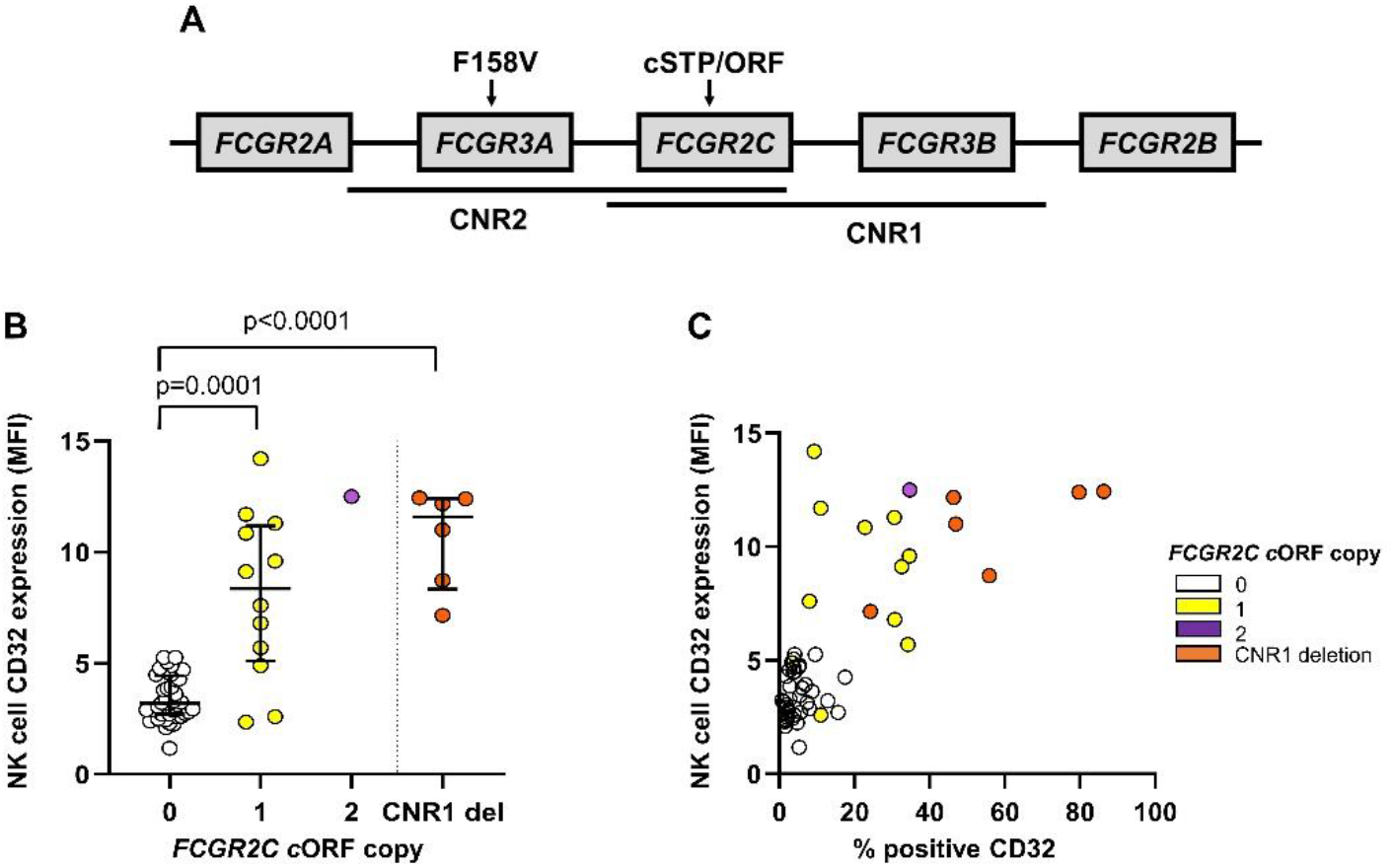
Novel *FCGR2C* QSV assay is associated with CD32 expression on natural killer cells. **(A)** Schematic of the two main copy number variable regions (CNVR1 and 2) and the relative positions of the functionally relevant nonsynonymous variants affecting *FCGR3A* and *FCGR2C*. **(B)** Expression of CD32 on natural killer (NK) cells in RA patients stratified by *FCGR2C* classical open reading frame (cORF) copy number. Using a novel *FCGR2C* QSV assay described in Supplementary Figure 2, in combination with multiplex ligation dependent assay probe values for rs10917661, the number of copies of the *FCGR2C* cORF were genotyped where matched NK cell CD32 (clone KB61) expression data were available. A particular locus rearrangement including a deletion of one copy of *FCGR2C* and one copy of *FCGR3B* (CNR1 del) and a STP allele on the remaining copy of *FCGR2C*, has been described to encode expression of FcγRIIb on NK cells. **(C)** The relationship between % positive CD32 expression on NK cells, and geometric mean fluorescence intensity (MFI). P-values calculated using non-parametric Mann-Whitney test. Data are summarized using median and interquartile range.

Using early RA patients prior to treatment, we observed increased NK cell CD32 expression in those with carriage of the classical *FCGR2C* ORF allele (p<0.0001, Figure 1B). Individuals carrying a deletion of CNR1 demonstrated high mean levels of NK cell CD32 expression (Figure 1B) with CD32 expression present on the majority of NK cells (Figure 1C), which suggests FcγIIb was expressed on these NK cells. Therefore, our results indicate that detailed copy number and SNP allele data are required to determine whether patient NK cells express either an activatory (FcγIIc) or inhibitory (FcγIIb) class II FcγR.

### Association of FCGR genotype and copy number with rituximab clinical response in RA

We next analysed the associations of *FCGR* genetic variants and gene and allele copy number with clinical response and depth of B cell depletion in RA and SLE. While the clinical variables predicting these outcomes were quite variable, the *FCGR* data were remarkably consistent.

Following quality control, we retained genotype data on 481 RA patients. Two SNPs in *FCGR2A* and one in *FCGR2B* were analysed in relation to clinical response (Table 3). In *FCGR2A*, for rs1801274 (H131R), there was weak evidence of a better outcome associated with the R allele and the 3-component DAS (p=0.04, not adjusted for multiple testing), with an inferior response for individuals homozygous for the H allele (p=0.03), but no evidence to support an association between rs9427399 (Q27W) or SNP I123T (rs1050501) in *FCGR2B* with any clinical response measures. The rs396991 (F158V) variant in *FCGR3A* was first analysed at the genotypic level with some evidence that the V allele was associated with a better response, particularly for SJC and 2C-DAS28CRP Table 3); carriers of at least one V allele had 5 fewer swollen joints than those with only F alleles (p=0.01) and 2C-DAS28CRP was on average 0.29 units lower than those with only F alleles (p=0.02), after adjusting for baseline measures. Compared to patients with two copies of *FCGR3A* (the majority), those with duplications were found to have a better clinical response on the basis of 2C-DAS28CRP (p=0.03). There was also a suggestion that those with deletions had a worse outcome, although numbers were very small (n=9). These facets were combined by examining the effect of the number of copies of the V allele. Higher numbers of the *FCGR3A*-158V allele were associated with better response in the form of SJC (p=0.01) and 2C-DAS28CRP (p=0.02). In a model including both the number of copies of the V allele and the number of copies of the F allele, there was some evidence that copy number (or number of copies of the F allele) also contributed to response when the 2-component DAS was evaluated (p=0.04).

**Table 3.** Effect of *FCGR2A, FCGR3A, FCGR2C, FCGR3B and FCGR2B* genotype and copy number on clinical response to rituximab in rheumatoid arthritis.

| *Gene*<sup>1</sup> | Alleles/<br>Copy<br>Number | Effect on CRP at 6<br>months: coefficient (SE),<br>*p*-value, N<sup>2</sup> | Effect on SJC at 6 months:<br>coefficient (SE), *p*-value,<br>N<sup>2</sup> | Effect on DAS-2C at 6<br>months: coefficient (SE),<br>*p*-value, N<sup>2</sup> | Effect on DAS-3C at 6<br>months: coefficient (SE),<br>*p*-value, N<sup>2</sup> |
| --- | --- | --- | --- | --- | --- |
| **Genotypic Analyses** |  |  |  |  |  |
| *FCGR2A*<br>(Q27W) | Q<br>(Reference) | -<br>366 | -<br>351 | -<br>318 | -<br>316 |
|  | QW | 0.06 (0.10), 0.52, 110 | -0.07 (0.12), 0.58, 101 | -0.01 (0.15), 0.97, 92 | 0.11 (0.14), 0.44, 92 |
|  | W | 0.21 (0.40), 0.61, 5 | 0.31 (0.49), 0.52, 5 | 0.51 (0.57), 0.38, 5 | -0.15 (0.48), 0.76, 5 |
| *FCGR2A*<br>(H131R) | R<br>(Reference) | -<br>121 | -<br>115 | -<br>103 | -<br>102 |
|  | RH | -0.09 (0.10), 0.35, 247 | -0.01 (0.12), 0.93, 230 | -0.18 (0.15), 0.25, 212 | -0.08 (0.14), 0.59, 212 |
|  | H | 0.17 (0.12), 0.14, 113 | 0.12 (0.14), 0.40, 112 | 0.21 (0.18), 0.24, 100 | 0.34 (0.16), **0.03**, 99 |
| *FCGR3A*<sup>3</sup><br>(F158V) | F<br>(Reference) | -<br>204 | -<br>190 | -<br>175 | -<br>174 |
|  | FV | -0.03 (0.09), 0.73, 225 | -0.25 (0.11), **0.02**, 217 | -0.29 (0.13), **0.03**, 193 | -0.08 (0.12), 0.53, 192 |
|  | V | -0.03 (0.14), 0.81, 52 | -0.26 (0.17), 0.13, 50 | -0.28 (0.21), 0.17, 47 | -0.21 (0.19), 0.27, 47 |
| *FCGR2C*<sup>3</sup><br>(STP/ORF) | STP<br>(Reference) | -<br>337 | -<br>315 | -<br>289 | -<br>287 |
|  | STPORF | 0.11 (0.10), 0.27, 110 | -0.13 (0.12), 0.28, 105 | -0.02 (0.15), 0.89, 95 | 0.00 (0.14), 0.99, 95 |
|  | ORF | -0.08 (0.23), 0.73, 16 | -0.16 (0.30), 0.59, 14 | -0.27 (0.35), 0.45, 14 | -0.40 (0.32), 0.21, 14 |

| *Gene*<sup>1</sup> | Alleles/<br>Copy<br>Number | Effect on CRP at 6<br>months: coefficient (SE),<br>*p-value*, N<sup>2</sup> | Effect on SJC at 6 months:<br>coefficient (SE), *p-value*,<br>N<sup>2</sup> | Effect on DAS-2C at 6<br>months: coefficient (SE),<br>*p-value*, N<sup>2</sup> | Effect on DAS-3C at 6<br>months: coefficient (SE),<br>*p-value*, N<sup>2</sup> |
| --- | --- | --- | --- | --- | --- |
| *FCGR3B*<sup>3</sup><br>(NA1/NA2<br>haplotype) | NA2<br>(Reference) | -<br>202 | -<br>186 | -<br>172 | -<br>171 |
|  | NA2NA1 | 0.01 (0.09), 0.88, 196 | 0.18 (0.11), 0.12, 184 | 0.19 (0.14), 0.19, 166 | 0.26 (0.18), 0.52, 166 |
|  | NA1 | 0.09 (0.13), 0.48, 63 | -0.10 (0.16), 0.53, 64 | -0.02 (0.19), 0.91, 58 | 0.12 (0.18), 0.52, 57 |
| *FCGR2B*<br>(I123T) | I<br>(Reference) | -<br>372 | -<br>359 | -<br>327 | -<br>325 |
|  | IT | 0.02 (0.10), 0.85, 100 | 0.06 (0.13), 0.64, 89 | 0.05 (0.16), 0.75, 80 | 0.11 (0.14), 0.44, 80 |
|  | T | 0.42 (0.34), 0.22, 7 | -0.32 (0.45), 0.48, 6 | -0.37 (0.53), 0.49, 6 | -0.15 (0.48), 0.76, 6 |
| **Copy Number Analyses** |  |  |  |  |  |
| *FCGR3A*<br>(F158V) | 2 copies<br>(reference) | -<br>434 | -<br>412 | -<br>375 | -<br>373 |
|  | <2 copies | 0.18 (0.29), 0.52, 10 | 0.59 (0.34), 0.08, 10 | 0.83 (0.43), **0.05**, 9 | 0.47 (0.39), 0.23, 9 |
|  | >2 copies | -0.19 (0.15), 0.21, 37 | -0.36 (0.19), 0.06, 35 | -0.52 (0.24), **0.03**, 31 | -0.31 (0.22), 0.16, 31 |
|  | No. copies of<br>V allele | -0.02 (0.06), 0.77, 481 | -0.18 (0.07), **0.01**, 457 | -0.20 (0.09), **0.02**, 415 | -0.09 (0.08), 0.26, 413 |
| *FCGR2C*<br>(STP/ORF) | 2 copies<br>(reference) | -<br>362 | -<br>342 | -<br>312 | -<br>311 |
|  | <2 copies | 0.04 (0.15), 0.79, 42 | -0.06 (0.19), 0.73, 38 | -0.06 (0.22), 0.80, 36 | -0.03 (0.21), 0.88, 35 |
|  | >2 copies | -0.19 (0.13), 0.12, 59 | 0.01 (0.16), 0.95, 54 | -0.23 (0.19), 0.25, 50 | -0.26 (0.18), 0.15, 50 |
|  | No. copies of<br>ORF allele | 0.04 (0.07), 0.57, 463 | -0.11 (0.08), 0.18, 434 | -0.09 (0.10), 0.36, 398 | -0.09 (0.09), 0.31, 396 |
| *FCGR3B*<br>(NA1/NA2<br>haplotype) | 2 copies<br>(reference) | -<br>385 | -<br>370 | -<br>334 | -<br>333 |
|  | <2 copies | 0.03 (0.14), 0.84, 47 | -0.20 (0.17), 0.26, 43 | -0.12 (0.21), 0.57, 40 | -0.02 (0.20), 0.91, 39 |
|  | >2 copies | -0.09 (0.14), 0.49, 49 | 0.07 (0.17), 0.70, 44 | -0.02 (0.21), 0.92, 41 | -0.02 (0.19), 0.90, 41 |
|  | No. copies of<br>NA1 allele | 0.02 (0.06), 0.74, 461 | 0.06 (0.07), 0.46, 434 | 0.08 (0.09), 0.38, 396 | 0.13 (0.08), 0.13, 394 |

<sup>1</sup>Genes presented in chromosomal order on 1q23 centromere to telomere.
<sup>2</sup>Coefficient, standard error (SE) and p-value for the effect of the indicated genotype or copy number on outcome at 6-months compared with baseline genotype or copy number. Positive coefficients for clinical response outcomes indicate a worse outcome.
<sup>3</sup>*FCGR3A*, *FCGR2C* and *FCGR3B* are subject to copy number variation, analyses were performed according to biallelic genotype whereby the effect of heterozygosity and homozygosity for the rare allele were compared with homozygosity for the common allele.
N: number

Integrated copy number and functional SNP analysis was also used to analyse quantitative and qualitative *FCGR2C* and *FCGR3B* variation. There was no association between either classical STP/ORF polymorphism (rs10917661) in *FCGR2C* or the NA1/NA2 haplotype in *FCGR3B* and clinical response at the genotypic level, nor after accounting for CNV. Of the 54 individuals with a *FCGR3B* deletion, 52 also had a deletion of *FCGR2C*, suggesting a CNR1 deletion (Figure 1A) in the majority of these individuals. Two individuals had a deletion of *FCGR2C* and *FCGR3A* (CNR2).

### Association of FCGR genotype and copy number with rituximab clinical response in SLE

To provide cross-disease replication of our findings in RA, we performed an equivalent analysis between *FCGR* genetic variation and SLE clinical response measures Table 4). For rs1801274 (H131R), homozygosity for the H allele of *FCGR2A* was associated with an increased odds of BILAG MCR at 6 months (p=0.02), however, there was no effect of *FCGR2A* rs9427399 (Q27W) on clinical response, nor I123T (rs1050501) in *FCGR2B*. Concordant with our findings in RA, the rs396991 (F158V) variant in *FCGR3A* was associated with increased odds of BILAG MCR when analysed at the genotypic level, with homozygosity for the V allele demonstrating a twofold improved response (p=0.03). Carriage of at least one copy of the *FCGR3A*-158V allele was associated with a 1.8 fold improvement in odds of BILAG response (p=0.04) and 1.9 fold improvement in odds of BILAG MCR (p=0.02). In contrast to RA, we did not find any evidence of an association between *FCGR3A* copy number and clinical response, most likely because the majority of patients carried two copies of *FCGR3A* (248/262). For each copy of *FCGR3A* V allele there were increased odds of BILAG MCR at 6 months (p=0.01). At the genotypic level, the *FCGR2C* classical STP/ORF genotype was associated with 2 fold increased odds of BILAG MCR at 6 months (p=0.02), with carriage of the ORF allele being associated with a 2.2 fold improvement in odds of BILAG MCR (p=0.02). Furthermore, duplications of *FCGR2C* had a 3.1 fold increased odds of BILAG MCR at 6 months (p=0.02), and a 2 fold improved response per copy of the *FCGR2C* ORF allele (p=0.02). For *FCGR3B*, patients with duplications also had the highest odds of achieving BILAG MCR compared to those with two copies (p=0.01). These associations with *FCGR2C* and *FCGR3B* copy number may not be independent as 17/22 subjects with a *FCGR3B* duplication also had a *FCGR2C* duplication indicating strong linkage disequilibrium between the genes. However, we found no evidence of association between other *FCGR3B* allelic variation and SLE clinical response, which may indicate that the principle association signal arises from *FCGR2C*. Sixty eight SLE patients had deletions affecting *FCGR3B*, 62 of which coincided with a deletion of *FCGR2C* (CNR1) in these individuals. The remaining 6 *FCGR2C* deletions coincided with *FCGR3A* deletion (CNR2)

**Table 4.** Effect of *FCGR2A, FCGR3A, FCGR2C, FCGR3B and FCGR2B* genotype and copy number on clinical response to rituximab in systemic lupus erythematosus.

| *Gene*<sup>1</sup> | Alleles/<br>Copy Number | BILAG Response (Major or Partial Clinical<br>Response) at 6 months:<br>OR (95% CI), *p*-value, N<sup>2</sup> | BILAG Major Clinical Response at 6 months:<br>OR (95% CI), *p*-value, N<sup>2</sup> |
| --- | --- | --- | --- |
| **Genotypic Analyses** |  |  |  |
| *FCGR2A*<br>(Q27W) | Q<br>(Reference) | -<br>208 | -<br>208 |
|  | QW | 2.01 (0.95 - 4.27), 0.07, 48 | 1.67 (0.88 - 3.18), 0.12, 48 |
|  | W | 0.53 (0.07 - 3.84), 0.53, 4 | 2.15 (0.30 - 15.61), 0.45, 4 |
| *FCGR2A*<br>(H131R) | R<br>(Reference) | -<br>70 | -<br>70 |
|  | RH | 0.74 (0.40 - 1.38), 0.35, 134 | 1.14 (0.61 - 2.15), 0.68, 134 |
|  | H | 1.23 (0.56 - 2.68), 0.60, 58 | 2.33 (1.12 - 4.85), **0.02**, 58 |
| *FCGR3A*<sup>3</sup><br>(F158V) | F<br>(Reference) | -<br>126 | -<br>126 |
|  | FV | 1.93 (1.10 - 3.39), **0.02**, 109 | 1.76 (1.02 - 3.06), **0.04**, 109 |
|  | V | 1.27 (0.53 - 3.06), 0.59, 27 | 2.51 (1.07 - 5.89), **0.03**, 27 |
| *FCGR2C*<sup>3</sup><br>(STP/ORF) | STP<br>(Reference) | -<br>173 | -<br>173 |
|  | STPORF | 2.20 (0.96 - 5.05), 0.06, 42 | 2.24 (1.13 - 4.43), **0.02**, 42 |
|  | ORF | 0.78 (0.13 - 4.78), 0.79, 5 | 1.36 (0.22 - 8.35), 0.74, 5 |
| *FCGR3B*<sup>3</sup><br>(NA1/NA2<br>haplotype) | NA2<br>(Reference) | -<br>106 | -<br>106 |
|  | NA2NA1 | 1.03 (0.58 - 1.81), 0.93, 111 | 1.01 (0.58 - 1.79), 0.96, 111 |
|  | NA1 | 1.23 (0.56 - 2.70), 0.60, 42 | 1.52 (0.73 - 3.17), 0.26, 42 |
| *FCGR2B*<br>(I123T) | I<br>(Reference) | -<br>197 | -<br>197 |
|  | IT | 0.68 (0.37 - 1.23), 0.20, 61 | 0.84 (0.45 - 1.55), 0.58, 61 |
|  | T | 1.31 (0.13 - 12.89), 0.82, 4 | 1.86 (0.26 - 13.46), 0.54, 4 |
| **Copy Number Analyses** |  |  |  |
| *FCGR3A*<br>(F158V) | 2 copies (reference) | -<br>248 | -<br>248 |
|  | <2 copies | See Note<sup>4</sup>, 5 | 0.46 (0.05 – 4.20), 0.49, 5 |
|  | >2 copies | 0.61 (0.16 – 2.32), 0.47, 9 | 0.53 (0.11 – 2.60), 0.43, 9 |
|  | No. copies of V<br>allele | 1.38 (0.92 – 2.06), 0.12, 262 | 1.64 (1.12 – 2.41), **0.01**, 262 |
| *FCGR2C*<br>(STP/ORF) | 2 copies (reference) | -<br>173 | -<br>173 |
|  | <2 copies | 1.43 (0.53 – 3.82), 0.48, 23 | 1.92 (0.80 – 4.61), 0.15, 23 |
|  | >2 copies | 1.71 (0.60 – 4.88), 0.31, 22 | 3.02 (1.22 – 7.48), **0.02**, 22 |
|  | No. copies of ORF<br>allele | 1.55 (0.80 – 3.00), 0.19, 220 | 1.93 (1.09 – 3.42), **0.02**, 220 |
| *FCGR3B*<br>(NA1/NA2<br>haplotype) | 2 copies (reference) | -<br>212 | -<br>212 |
|  | <2 copies | 1.22 (0.51 – 2.93), 0.65, 27 | 1.52 (0.67 – 3.46), 0.32, 27 |
|  | >2 copies | 2.31 (0.76 – 7.09), 0.14, 22 | 3.20 (1.30 – 7.85), **0.01**, 22 |
|  | No. copies of NA1<br>allele | 1.18 (0.81 – 1.72), 0.39, 259 | 1.24 (0.86 – 1.78), 0.24, 259 |

<sup>1</sup>Genes presented in chromosomal order on 1q23 centromere to telomere.
<sup>2</sup> Odds ratio (OR), 95% confidence intervals (CI) and *p*-value for the effect of the indicated copy number on outcome at 6 months. All tests were performed using univariable logistic regression.
<sup>3</sup>*FCGR3A*, *FCGR2C* and *FCGR3B* are subject to copy number variation, analyses were performed according to biallelic genotype whereby the effect of heterozygosity and homozygosity for the rare allele were compared with homozygosity for the common allele.
<sup>4</sup>All 5 individuals with a deletion achieved British Isles Lupus Assessment Group (BILAG) any response, so the OR was not estimated as the outcome was predicted perfectly
N: number

There were insufficient individuals from non-Caucasian backgrounds with 2 copies of *FCGR3A*, *FCGR2C* and *FCGR3B* to accurately determine whether this group had different patterns of linkage disequilibrium to the Caucasian population. A sensitivity analysis was therefore performed in Caucasians only, with broadly similar results, whereby the per allele effect size of major clinical response was 1.62 (0.97 – 2.71, p = 0.07) for *FCGR3A*-158V and 1.75 (0.91 – 3.36, p = 0.09) for *FCGR2C*-ORF (Supplementary Table 1)

### Association of FCGR F158V genotype and copy number with complete B cell depletion in RA and SLE

Since peripheral B cells were analysed using HSFC in the Leeds cohorts only, data from both RA and SLE were combined to increase statistical power (n=413). There was no significant difference in depth of depletion between the disease groups (p=0.26), although there was a significant difference in age (p=0.002). The baseline clinical variables associated with depletion in the combined group are shown in Supplementary Table 2. Older age (p=0.02), concomitant DMARDs with (p=0.02), or without including HCQ (p=0.003), IgA (p=0.01), IgG (p=0.003) and plasmablasts (p<0.001) were all associated with complete B cell depletion. A significant correlation between IgG and baseline plasmablast counts was observed (p=0.01).

Downstream genetic analyses are presented both unadjusted and adjusted for age, concomitant DMARDs including HCQ and plasmablasts (Figure 2; Supplementary Table 3). Of the two genes unaffected by CNV (*FCGR2A* and *FCGR2B*), no association with complete B cell depletion was observed. For the CNV affected genes, higher number of copies of the *FCGR3A* V allele were associated with an increased odds of B cell depletion (p=0.02), in both the adjusted and unadjusted analyses.

**Figure 2:**
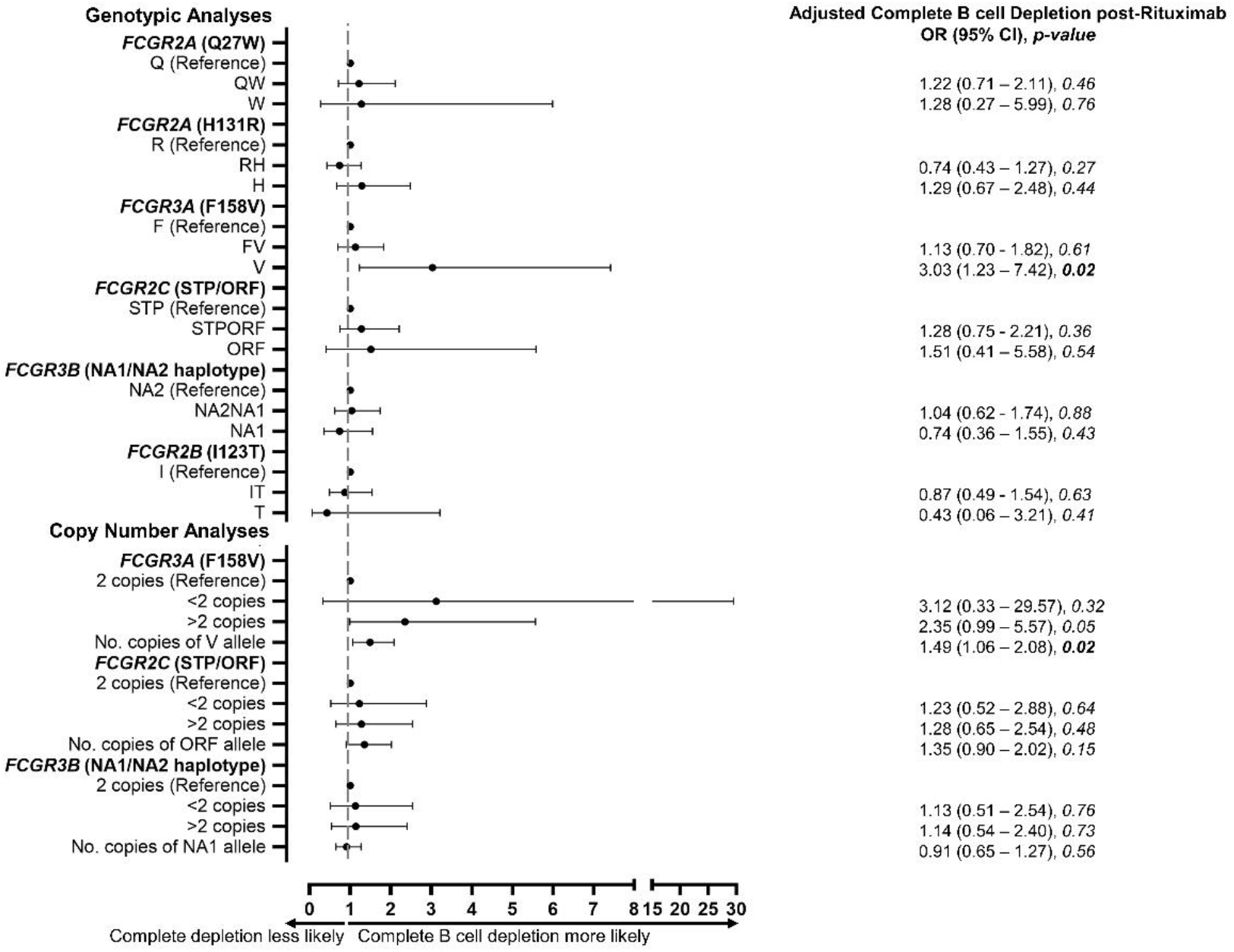
Association of *FCGR* genotype and copy number with complete B cell depletion. Odds ratio (OR), 95% confidence intervals (CI) and p-value for the effect of the indicated genotype or copy number on complete B-cell depletion at 2 weeks post-rituximab, compared with reference genotype. The dots represent the OR and the error bars denote the CI. All tests were performed using univariable logistic regression, adjusted for age, concomitant disease-modifying anti-rheumatic drug, including hydroxychloroquine, and baseline plasmablast count.

### Characterisation of NK cells, FcγR expression and FcγR effector functions in RA and SLE

*FCGR3A* genetic variation may impact clinical responses, including B cell depletion, through increased expression of FcγRIIIa on the cell surface and/or altered rituximab-induced ADCC. To disentangle gene and disease-specific effects on NK cell number and function, we functionally characterised NK cell FcγRIIIa expression and cellular cytotoxicity in healthy controls (HC), RA and SLE patients.

### Factors that modulate NK cell FcγRIIIa expression in the RA disease continuum and SLE

NK cell FcγRIIIa expression was measured by flow cytometry in HC (n=47), early RA (symptom onset <1 year and DMARD-naïve; n=46) and established RA (diagnosed >2 years and receiving DMARDs; n=20). FcγRIIIa expression differed between the three groups (p=0.01) and was reduced in early RA (p<0.001), but not established RA (p=0.22), when compared to HC Figure 3A). This differential expression was not secondary to altered ratios of CD56^bright^ to CD56^dim^ NK cells between RA patients and HC Supplementary Figure 1A). Early RA patients showed lower FcγIIIa expression on both NK cell subsets (both p=0.02) compared to HC (Supplementary Figure 1B and 1C). Indeed, there was no difference in FcγIIIa expression between HC and established SLE patients recruited on the day of their first rituximab cycle Supplementary Figure 1D).

**Figure 3.**
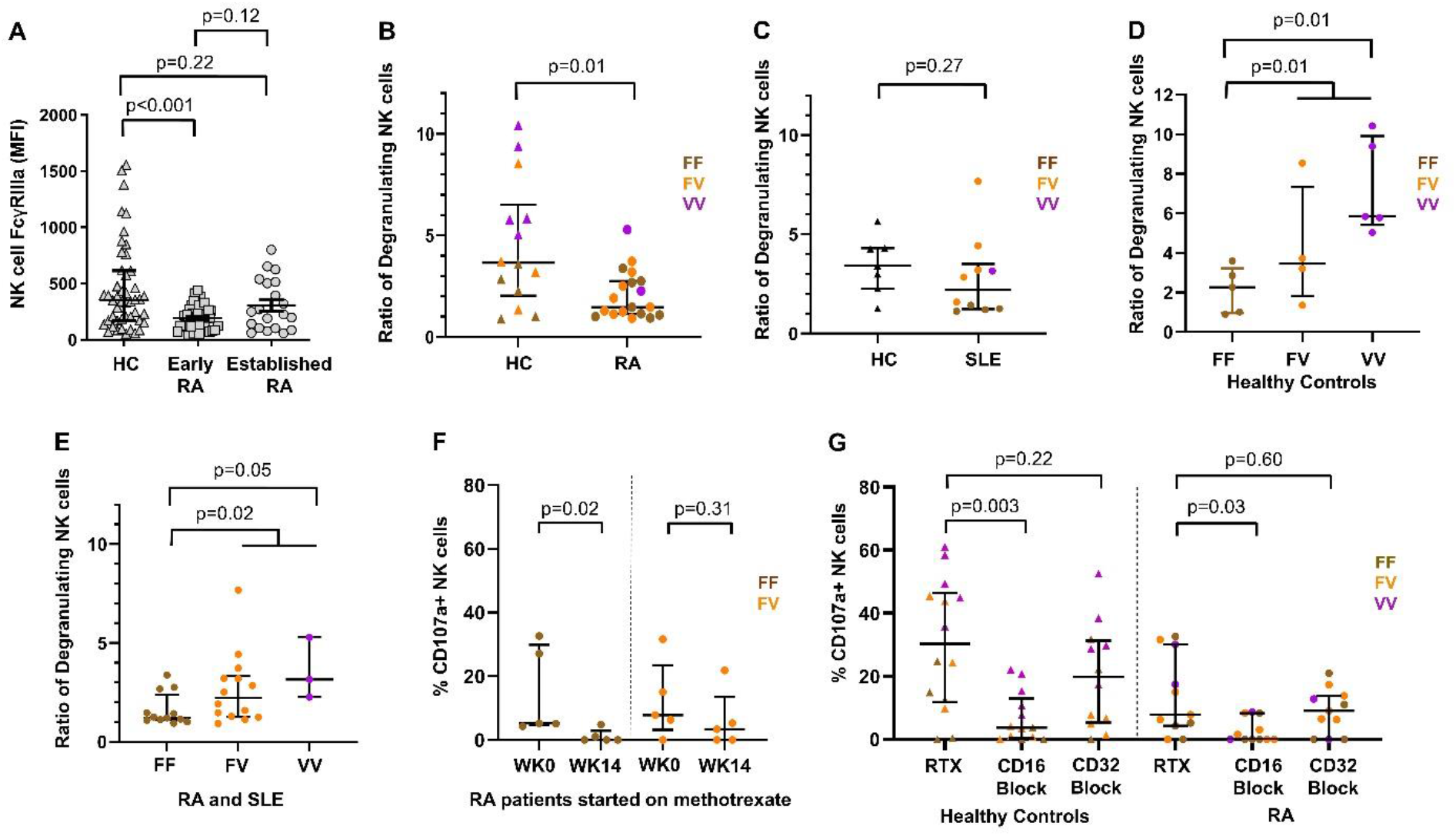
Effect of disease, genotype and methotrexate on natural killer cell degranulation. **(A)** Natural killer (NK) cell (CD3-CD56+CD16+) FcγRIIIa (CD16; clone 3G8) geometric mean fluorescence intensity (MFI) using flow cytometry for healthy controls (HC), early (symptom onset <1 year and treatment naïve) and established (>2 years) rheumatoid arthritis (RA). Comparison of NK cell degranulation following incubation with B cell lineage (Daudi cells for RA **(B)** and Raji cells for systemic lupus erythematosus (SLE; **C)**) using E:T ratio of 1:1 and rituximab. Ratio of degranulating NK cells were compared in individuals with two copies of *FCGR3A* between the three *F158V* genotypes in HC **(D)** and RA and SLE combined **(E)**. **(F)** NK cell degranulation (%CD107a positive NK cells) was compared before and 14 weeks after RA patients started on methotrexate according to their *FCGR3A* genotype. **(G)** %CD107a positive NK cells was assessed after incubation with rituximab and subsequent inclusion of CD16 (clone 3G8) and CD32 (clone KB61) blocking antibodies in HC and RA.

To assess factors contributing to the downregulation of NK cell FcγRIIIa expression in early RA, we examined its relationships with clinical and serological markers. There were moderate correlations between NK FcγRIIIa expression and RF titre (r=0.38; p=0.03) and serum IL-6 titre (r=0.38; p=0.03), but not CRP (r=0.14; p=0.47), ESR (r=0.16; p=0.38), DAS28 (r=-0.09; p=0.69), age (r=-0.02; p=0.92), nor gender (p=0.82).

NK FcγRIIIa expression was then examined in relation to the *FCGR3A*-F158V variant in individuals with 2 copies of *FCGR3A*. No significant differences in NK FcγRIIIa expression were demonstrated between the *FCGR3A* genotypic groups in either HC (p=0.74; Supplementary Figure 1E), RA (p=0.96; Supplementary Figure 1F) or SLE (p=0.21; Supplementary Figure 1G).

### Disease, genotype and DMARDs impact efficiency of NK cell-mediated ADCC

NK cell degranulation (CD107a staining) following exposure to rituximab-coated B lineage cell lines was used as a surrogate of ADCC in individuals with 2 copies of *FCGR3A*. NK cell degranulation was significantly decreased in RA (p=0.01; Figure 3B), but not SLE (p=0.27; Figure 3C) patients compared with HC. *FCGR3A* genotype was associated with rituximab-induced NK cell degranulation in HC, RA and SLE patients, whereby carriage of the 158V allele and 158V homozygotes were associated with improved degranulation in HC (both p=0.01; Figure 3D), and in RA and SLE (p=0.02, 0.05; Figure 3E), respectively, compared to non-carriers.

The reduced degranulation in RA compared with HC, prompted us to explore the impact of DMARDs on NK cell degranulation *ex vivo* (Figure 3F). A significant reduction in the number of NK cells degranulating was observed after 14 weeks of methotrexate in RA, compared to baseline, with the most marked reduction occurring in individuals homozygous for the *FCGR3A-158F* allele (p=0.02).

Inclusion of a CD16 blocking antibody inhibited rituximab-induced NK cell degranulation in HC (p=0.003) and RA (p=0.03, Figure 3G), and thus supported a major role for FcγRIIIa in NK cell-mediated ADCC *ex vivo*. No significant reduction in NK cell degranulation was observed with CD32 blockade in HC or RA (p=0.22, p=0.60; Figure 3G). None of these individuals had a deletion in CNR1 (FcγRIIb expression) and there was no clear association with *FCGR2C* genotype.

### Higher expression of FcγRIIIa is associated with better clinical response and depletion in vivo independent of NK cell number

We measured absolute NK cell numbers and NK cell FcγRIIIa expression on the day of the first infusion of rituximab compared with subsequent depletion and response to explore these effector mechanisms *in vivo*.

Absolute NK cell number was lower in SLE than RA (p<0.001; Figure 4A). However, there was no evidence for an association between NK cell number and EULAR response (p=0.80; Figure 4B) or complete B cell depletion (p=0.68; Figure 4C) in RA, nor BILAG MCR (p=0.96; Figure 4D) or complete B cell depletion (p=0.67; Figure 4E) in SLE.

**Figure 4.**
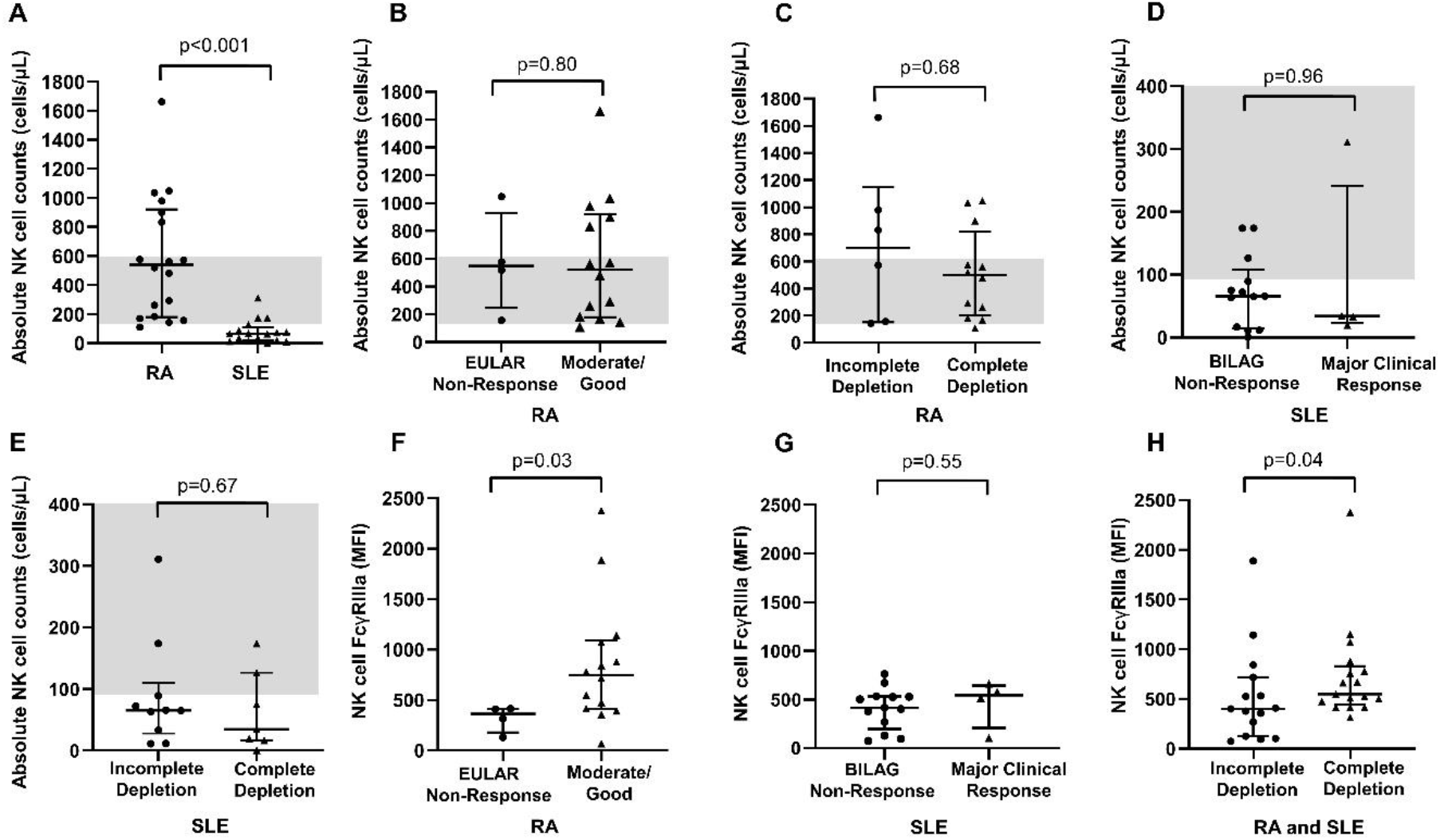
Peripheral blood natural killer cell abundance and FcγRIIIa expression in rheumatoid arthritis and systemic lupus erythematosus patients treated with rituximab. Comparison of absolute natural killer (NK) cell (CD3-CD56+CD16+) counts between **(A)** rituximab-treated rheumatoid arthritis (RA) and systemic lupus erythematosus (SLE) patients; **(B)** rituximab-treated RA patients exhibiting no European League Against Rheumatism (EULAR) clinical response and moderate/good clinical response; (**C**) rituximab-treated RA patients exhibiting incomplete and complete B cell depletion; **(D)** rituximab-treated SLE patients exhibiting no clinical response and major clinical British Isles Lupus Assessment Group (BILAG) response; and **(E)** rituximab-treated SLE patients with incomplete and complete B cell depletion. The shaded grey areas represent adult reference ranges (90-600 cells/µL). Expression of FcγRIIIa (CD16; clone 3G8) on NK cells of **(F)** RA patients exhibiting EULAR non-response and moderate/good clinical response to rituximab; **(G)** SLE patients exhibiting BILAG non-response and major clinical response to rituximab; and **(H)** RA and SLE patients exhibiting incomplete and complete B cell depletion in response to rituximab.

We measured NK cell FcγRIIIa expression in 18 RA patients and 17 SLE patients immediately prior to rituximab and demonstrated a positive association between FcγRIIIa expression and EULAR response at 6 months (defined as improvement of at least 0.6 point to DAS28CRP≤5.1) in RA (p=0.03; Figure 4F), but not BILAG MCR in SLE (p=0.55, Figure 4G). There was no difference in FcγRIIIa expression between RA and SLE (p=0.14) and the groups were combined to explore the effect of FcγRIIIa expression on B cell depletion. Patients with complete depletion had higher NK cell FcγRIIIa expression at rituximab initiation than those with incomplete depletion in RA and SLE (p=0.04: Figure 4H).

## DISCUSSION

We report the largest study, to date, of quantitative and qualitative functional variants at the *FCGR* genetic locus in well-characterised RA and SLE cohorts, including subgroups where peripheral blood B cell (naive, memory and plasmablast cell) levels were available before and after rituximab therapy. We provide consistent evidence that FcγRIIIa is the major low affinity FcγR associated with both clinical and biological (i.e. depth of B cell depletion) response to rituximab in both autoimmune diseases. More specifically, an increase in the number of copies of the *FCGR3A*-158V allele, encoding the allotype with a higher affinity for IgG1, was associated with improved responses. In addition, irrespective of *FCGR3A* genotype, we observed that the higher the number of *FCGR3A* gene copies correlated with a better clinical response in RA. To functionally support these genetic findings, we also demonstrated that *FCGR3A* genotype was associated with NK cell-mediated degranulation *in vitro*, and increased NK cell FcγRIIIa expression was associated with improved clinical response and depletion *in vivo*, thus providing further biological support for our genetic studies.

In RA, we have presented data on the commonly used clinical endpoints of CRP, SJC and 3C-DAS28CRP. We also included our more recently published 2C-DAS28CRP score (*35*) that includes revised weighting of CRP and SJC to more closely reflect the more objective endpoint of ultrasound-proven synovitis, an outcome measure that we have proposed as the RA disease activity measure of choice for genetic and biomarker studies (*36*). In SLE we have used a BILAG-based endpoint rather than SLEDAI based. BILAG is better for biomarker studies because, unlike the SLEDAI, it allows partial improvement, it allows severe and moderate manifestations within each organ system to be weighted equally, and it does not include serological components – a serious confounder in studies of a B cell-targeted therapy.

Our studies have once again highlighted the challenges of analysing and interpreting the *FCGR* genetic locus, even with commercially available assays. We present the first study that has undertaken a combined quantitative and qualitative analysis of the *FCGR* locus to allow more comprehensive biological interpretation. The association with *FCGR3A* was most apparent when analysed as the number of copies of the 158V allele, consistent with previous studies in RA where traditional SNP analyses demonstrated improved responses in those that carried the *FCGR3A-158V* allele (*27, 37*).

No consistent association signals were observed with other low affinity *FCGR* genes at this locus, suggesting FcγRIIIa was the most important FcγR contributing to rituximab response. Increased NK cell-mediated degranulation was observed in individuals homozygous for the *FCGR3A* V allele, irrespective of disease status, which combined with the association between FcγRIIIa expression on NK cells and *in vivo* clinical response, independent of absolute NK cell numbers, supports ADCC being a major biological mechanism. However, this does not preclude FcγRIIIa-mediated clearance of rituximab-opsonised B cells by tissue macrophages and cells of the reticuloendothelial system by ADCP (*38*). Our data also reveal potential disease or inflammation-specific factors that may impair ADCC, for example reduced NK cell degranulation and reduced FcγRIIIa expression in early RA patients compared with HC, which we showed was correlated with serum IL-6 titre and autoantibody status, but not *FCGR3A*-158F/V genotype.

Furthermore, we demonstrated reduced ADCC in RA patients after 14 weeks of MTX therapy compared with a pre-DMARD sample at diagnosis, suggesting that the medication used at the time of rituximab treatment may modulate NK cell-mediated effector mechanisms. Paradoxically, our clinical data demonstrated more robust responses in patients taking concurrent DMARDs, suggesting additional biological pathways may contribute to clinical outcomes. Further studies are now required to understand how NK cell function can be optimised at the time of rituximab infusions to improve clinical outcomes.

In SLE, higher number of copies of the *FCGR2C* ORF allele and *FCGR2C* gene duplications were also associated with BILAG MCR at 6 months. At the time this study was carried out, the commercial MLPA panels did not include probes for *FCGR2C* copy number *per se* and these panels were supplemented with additional *FCGR2C* genotyping to aid interpretation. Our expression studies demonstrated that NK cell CD32 expression correlated with carriage of the *FCGR2C*-ORF allele, but also revealed high expression levels in individuals with simultaneous deletion of *FCGR2C* and *FCGR3B*, which leads to FcγRIIb expression. This rearrangement was observed in 52 RA and 62 SLE individuals in our genetics studies, with no evidence it significantly impacted on depletion or response to therapy.

Our results have implications for the design of future clinical trials optimising rituximab therapy and development of more effective B cell targeted strategies, which can be extrapolated to other B cell-mediated autoimmune diseases. For example, patients with high copy number of *FCGR3A*-158V may be most suitable for stratification to lower doses of rituximab (i.e. 500mg x 2) or even ultra-low dose regimen (i.e. 200mg x 1) (*39*) during repeat rituximab cycles. This may reduce long-term complications, such as late onset neutropaenia (*31*) and infections secondary to hypogammaglobulinaemia (*40*). If successful this would reduce drug costs and administration times. A previous clinical trial has shown that patients with initial incomplete depletion can benefit from a higher dose of rituximab than currently licensed (1000mg x 3) (*41*). Entry into that study was determined after measuring initial depletion, but *FCGR3A* genotyping may allow stratification prior to administering treatment. Secondly, confirmation that FcγRIIIa is the major FcγR contributing to clinical response is of particular value to therapeutic antibody design and highlights the need for the next generation of CD20 therapeutic antibodies to show equivalent ADCC potency in individuals with both FcγRIIIa-158F/V allotypes, particularly when based on the native IgG1 background. Indeed, there are many novel therapeutics in development with modified Fc regions that enhance ADCC. Obinutuzumab was one of the first glycoengineered therapeutic mAbs to be FDA approved. Here control of fucosylation during manufacture leads to increased ADCC, albeit at the expense of reduced CDC (*42*). This type 2 CD20 mAb binds to a different CD20 epitope and is currently being investigated in a phase III trial of lupus nephritis [NCT04221477], with future trials planned in RA. Other approaches to optimising effector function through modification of the Fc include point mutations to the CH2 and CH3 domains, selected for their combined ADCC-enhancing effect for both FcγRIIIa-158F and -V (e.g. margetuximab), and asymmetrically engineered Fc domains through the mutation of different amino acids in each Fc domain (*43*). Thirdly, further consideration also needs to be given for factors determining FcγRIIIa expression and whether expression levels can be increased, or NK cell function optimised at the time of treatment. Finally, there are also implications for quality control of biosimilar rituximab, where significant batch-batch differences in ADCC are recognised and may disproportionally affect individuals homozygous for the *FCGR3A*-138F allele (*44*).

The challenge for pharmacogenetic studies is the assembly of large cohorts of sufficiently homogeneous patients. Efforts were made in this study to minimize heterogeneity by using strict entry criteria and improve consistency of time-points and definition of clinical response, however, some clinical and laboratory data were missing, which is inevitable when large observation cohorts are utilised. Most notably, B cell data were only available for the patient cohorts treated in Leeds. Hence, some analyses were performed by combining data from RA and SLE. We have presented data on a number of outcome measures to support future meta-analyses. For the RA cohort, 481 patients were genotyped. This sample size gives over 80% power to detect a genetic variant explaining 2.5% of the variance in outcome at a nominal significance level of 0.01, but a sample size of at least 1200 patients would be needed to have over 80% power to detect a variant explaining only 1% of the variance. We have presented nominal p-values without taking into account the number of analyses conducted, and p-values must be interpreted bearing this in mind. The strength of the evidence for the role of *FCGR3A* lies in the consistent direction of genetic association in two different diseases and by the *in vitro* and *in vivo* studies.

In conclusion, an ADCC-enhancing quantitative *FCGR3A* variant was associated with clinical response and complete B cell depletion in rituximab-treated patients with RA and SLE. These findings were supported by our mechanistic studies demonstrating the impact of FcγRIIIa expression, genotype, disease status and medication on NK cell-mediated cytotoxicity. These results elucidate one mechanism of impaired rituximab responses, may guide development of more effective B cell targeted strategies and emphasise the importance of ensuring the next generation of therapeutics bind with equivalent affinity to both FcγRIIIa allotypes.

## MATERIALS AND METHODS

### Study design

A mixture of prospective and retrospective longitudinal cohort studies were conducted in patients with RA and SLE who were treated with rituximab from January 2001 to January 2020. These patients were recruited from two UK biologic DMARD (bDMARD) registries and a large cohort from the Leeds Teaching Hospitals NHS Trust (LTHT), UK.

Full details of the patient cohorts, clinical outcome measures, treatment and routine laboratory assessments are provided in the **Supplementary Materials and Methods**.

### Ethical approval

All component studies were approved by a Research Ethics Committee (REC); North West Multicentre (00/8/053), North West Greater Manchester (09/H1014/64), COREC (04/Q1403/37), Leeds West (01/023, 05/MRE03/85 and 09/H1307/98) and Leeds East (04/Q1206/107, 10/H1306/88). All participants provided informed written consent and research was carried out in compliance with the Declaration of Helsinki. All experiments were performed in accordance with relevant protocols and regulations.

### Highly Sensitive Flow Cytometry

Peripheral blood B cell subsets were measured using HSFC at the accredited Leeds Haematological Malignancy Diagnostic Service at baseline and on follow-up, as previously described (*45*). A six colour flow cytometry protocol (CD3, CD14, CD19, CD27, CD38, CD45), counting 500,000 events was used. Naive (CD19+CD27-), memory (CD19++CD27+) and plasmablast (CD19+/-CD27++CD38++) counts were enumerated using CD45 to identify the total leucocyte population for calculation of absolute B cell subset numbers, using CD3 and CD14 to exclude contaminating leucocyte populations. Complete B cell depletion was defined as a total B cell count ≤0.0001 x 10^9^ cells/L at week 2 for RA and week 6 for SLE.

### Genotyping

Genomic DNA was extracted from EDTA-anticoagulated whole blood using Qiamp mini spin columns (Qiagen), Gene Catcher (ChargeSwitch, Thermo Fisher) and a manual phenol chloroform method. DNA concentration was measured using UV spectrophotometry and samples diluted to 10ng/µl. 50ng total template was used for each MLPA reaction. MLPA probe mix panels P110 and P111 (MRC-Holland, Amsterdam, The Netherlands) were performed for every sample. Where a sample failed in the first reaction due to insufficient template (Q fragment QC) the DNA template was concentrated using ethanol precipitation and MLPA repeated. MLPA panels were supplemented with an in-house assay which yielded an independent measurement of *FCGR2C* gene copy number by interrogating a unique *FCGR2C* specific nucleotide discriminating all three *FCGR2* genes from each other (**Supplementary Materials and Methods,** Supplementary Figure 2). Amplified MLPA fragments were separated against a LIZ GS 5500 size standard using an ABI 3130 Applied Biosystems (Warrington, UK), instrument fitted with a 36cm 16 capillary array. Intra-sample normalisation against internal amplification controls and reference probes was performed using Coffalyser.NET software (MRC-Holland), prior to inter sample normalisation in batch analysis mode. Interpretation of the underlying gene copy number and qualitative variants followed the guidelines set out in the product information sheets.

### Flow cytometry and NK cells

NH_4_Cl lysed heparinised blood samples for RA and PBMCs for SLE were incubated with 10μg/ml human IgG (Novartis, Hanover, Germany), to block non-specific binding, for 30min on ice. Cells were stained (30 min, on ice, in dark) with anti-CD16-PE (clone 3G8, Caltag-Medsystems, Buckingham, UK), anti-CD32 (clone KB61, Dako, Cambridge UK), anti-CD3-TR (S4.1, Caltag-Medsystems, Buckingham, UK) and anti-CD56-PerCP (B159, Becton Dickinson Biosciences, Berkshire, UK) or isotype controls for experiments in RA and matched HC, while cells were stained with anti-CD16-APC Vio770 (REA423, Miltenyi Biotec, Surrey, UK), anti-CD3-VioGreen (REA613, Miltenyi Biotec, Surrey, UK) and anti-CD56-VioBright FITC (AF12-7H3, Miltenyi Biotec, Surrey, UK) or isotype controls for experiments in SLE and matched HC. Cells were then washed in ice cold FACS Buffer and 5000 events were acquired within the appropriate forward scatter (FSC)/side scatter (SSC) and CD3-gates on FACSCalibur (BD Biosciences, Berkshire, UK). Peripheral blood NK cells (CD3-CD56+CD16+) were identified on the basis of lymphocyte FSC/SSC characteristics and specific gating for lack of expression of CD3 and positive expression of CD56 (all NK cells, Supplementary Figure 3A). The CD16/CD32 geometric mean fluorescence intensity (MFI) and the percentage of CD16/CD32 positive cells were determined. NK cell subsets were distinguished by separate gates created around CD56^dim^/CD16++ and CD56^bright^/CD16^neg/low^ NK cells (Supplementary Figure 3B; dashed and solid boxes, respectively).

Absolute NK numbers (cells/μL) = Lymphocyte count (cell number/μL of the blood count) x proportion of the cell subpopulation of interest ÷ 100. Adult reference ranges are 90-600 cells/µL (*46*).

### NK cell degranulation assays

#### Rheumatoid arthritis and matched controls

Pre-genotyped, thawed cryopreserved PBMCs isolated from NH_4_Cl-lysed heparinised blood using a Ficoll-Paque gradient were recovered overnight in RPMI Glutamax media (Invitrogen, Loughborough, UK) containing 10% FBS at a concentration of 2×10^6^ cells/ml in the presence of 400U/ml of IL-2 (Sigma, Dorset, UK). All experiments were performed at 37℃ in a humidified 5% CO_2_ incubator, unless otherwise stated. NK cell degranulation assays were performed over 4 hours in the presence of CD107a FITC antibody (H4A3; BD Biosciences, Berkshire, UK) using recovered PBMCs and human B lineage cell lines (Daudi cells; ATCC, Manassas, Virginia, USA) that had been pre-incubated with rituximab (Roche Ltd, Basel, Switzerland) at 10µg/ml overnight using effector-target (E:T) ratios of 1:1 (*47, 48*). GolgiStop™ (BD Biosciences, Berkshire, UK) was added after 1 hour to halt protein transport and prevent internalisation of CD107a. Negative controls included Daudi cells incubated overnight in the absence of rituximab (E:T of 1:1; spontaneous degranulation) and assays performed without CD107a in the presence of a FITC-isotype control (Dako, Santa Clara, USA). After 4 hours, cells were washed and stained with anti-CD3 (S4.1; Invitrogen, Loughborough, UK), anti-CD14 (MφP9; BD Biosciences, Berkshire, UK) and anti-CD56 (B159; BD Biosciences, Berkshire, UK) for 30mins (on ice, in dark) then washed in ice cold FACS buffer and flow cytometric analysis was performed as outlined above. The ratio of CD107a positive NK cells without:with rituximab, identified by FSC/SSC, CD3 and CD56 expression, was calculated using the Flowjo software analysis package (BD Biosciences, Berkshire, UK). Additional blocking assays, were performed in a subset of experiments with anti-CD16 (3G8, Invitrogen, Loughborough, UK), anti-CD32 (KB61, Dako, Santa Clara, USA) and mouse IgG (Invitrogen, Loughborough, UK).

#### Systemic Lupus Erythematosus and matched controls

For experiments performed on SLE cases and matched controls, fresh PBMCs were initially isolated using Greiner Bio-One Leucosep™ centrifuge tubes with porous barrier (Thermo Fisher Scientific, Loughborough, UK). NK cells were isolated by negative selection using an NK Cell Isolation Kit and Immunomagnetic (Miltenyi Biotec, Surrey, UK) and cytofluorometric selection of CD3-CD56+ NK cells using flow cytometry resulting in ≥90% purity.

NK cell degranulation assays were performed by the addition of Rituximab (N7049B13, Roche Ltd, Basel, Switzerland) to NK cells incubated in the presence of Raji cells (Sigma-Aldrich, Gillingham, UK) using E:T ratio of 1:1, as well as GolgiStop™ (BD Biosciences, Oxford, UK) for 4 hours. Following incubation, anti-CD107a-FITC (REA792, Miltenyi Biotec, Surrey, UK) was added and NK cells co-stained with anti-CD3 (REA613, Miltenyi Biotec, Surrey, UK), anti-CD56 (AF12-7H3, Miltenyi Biotec, Surrey, UK) and anti-CD16 (REA423, Miltenyi Biotec, Surrey, UK) antibodies. Degranulation activity was measured by the ratio between (percentage CD107a positivity Raji cells only) and (percentage CD107a positivity in rituximab-coated Raji cells) Supplementary Figure 4). Flow cytometry was performed using a Becton Dickinson (BD) LSRII or BD FACSCalibur flow cytometer for data acquisition and using a BD FACSDiva software for data analyses.

#### Statistical analyses

Associations between baseline demographic, clinical and laboratory variables and clinical response measures were assessed using linear regression of the 6 month measures, adjusting for the corresponding baseline measure for RA; logistic regression was used for SLE clinical outcomes. Complete B cell depletion post-rituximab for the combined RA and SLE cohort was assessed using logistic regression, without adjustment and following adjusting for age, concomitant DMARD use and baseline plasmablast count. Positive coefficients for clinical response outcomes or B cell levels indicate a worse outcome, but odds ratios greater than 1 for the analysis of SLE outcomes or complete B cell depletion indicate a better outcome (i.e. greater chance of complete depletion).

A number of putatively functional genetic variants across five genes in the *FCGR* locus were analysed for associations with clinical response at 6 months (linear for RA and logistic regression for SLE and complete B cell depletion post-rituximab). For three SNPs in two genes where no copy number variation was observed (*FCGR2A* and *FCGR2B*), associations between the SNPs and outcomes were assessed using standard genotypic tests, whereby individuals homozygous for the common allele served as a reference group. For genes affected by CNV (*FCGR2C*, *FCGR3A*, *FCGR3B*), three analyses were carried out. The first, emulating previous studies that did not take CNV into consideration and assuming 2 gene copies; the second compared those with normal copy number of 2 genes with individuals with deletions and individuals with duplications, irrespective of which alleles were carried; the third based on the number of copies of the minor allele and overall copy number. To determine whether the number of copies of the minor allele improves on the nested model with overall copy number only, a likelihood ratio test was performed. Since the RA and SLE cohorts were of mixed ethnicity, we assessed the potential for population stratification by measuring pairwise LD between the relevant functional polymorphisms in the *FCGR* locus for each sub group. We utilised Haploview to calculate r^2^ LD between biallelic markers in individuals with two copies of *FCGR3A* and *FCGR3B* (*49*).

Continuous variables were compared using Mann-Whitney test or Kruskal-Wallis H test, depending on data distribution and number of independent groups for comparison. Spearman’s test was used for all correlations. Associations between categorical variables were tested by Fisher’s exact test if expected number is ≤5, otherwise chi-squared tests were performed. All statistical analysis was performed using StataMP v.16 (StataCorp College Station, Texas, USA), SPSS v.26 (IBM Corp, Armonk, New York, USA) and GraphPad Prism v.8.3 (GraphPad Prism, La Jolla, California, USA) for Windows.

## Data Availability

All data associated with this study are available in the main text or the supplementary materials. Anonymised data and statistical codes can be shared and all requests should be made to AWM to ensure all users of the data adhere to the legal requirements and data sharing of personal data. This work is licensed under a Creative Commons Attribution 4.0 International (CC BY 4.0) license, which permits unrestricted use, distribution, and reproduction in any medium, provided that the original work is properly cited. To view a copy of this license, visit http://creativecommons.org/licenses/by/4.0/.

## Acknowledgements

We thank all the patients who have contributed to this research, clinical staff who supported patient recruitment and laboratory staff who undertook sample processing.

AWM, INB, JDI and PE were supported by NIHR Senior Investigator awards. Work in JDI’s laboratory is supported by the NIHR Newcastle BRC, the Research Into Inflammatory Arthritis Centre Versus Arthritis, and Rheuma Tolerance for Cure (European Union Innovative Medicines Initiative 2, grant number 777357). INB is funded by the NIHR Manchester BRC.

This article/paper/report presents independent research funded/supported by the NIHR Leeds BRC and the NIHR Guy’s and St Thomas’ BRC. The views expressed are those of the author(s) and not necessarily those of the NIHR or the Department of Health and Social Care.

## Funding

We thank the Medical Research Council (MRC) and Versus Arthritis for their joint funding of MATURA (grant codes 36661 and MR/K015346/1). MASTERPLANS was funded by the MRC (grant code MR/M01665X/1). The Leeds Biologics Cohort was part funded by programme grants from Versus Arthritis (grant codes 18475 and 18387), the National Institute for Health Research (NIHR) Leeds Biomedical Research Centre (BRC) and Diagnostic Evaluation Co-operative and the Ann Wilks Charitable Foundation. The BILAG-BR has received funding support from Lupus UK, and unrestricted grants from Roche and GSK.

The functional studies were in part supported through a NIHR/HEFCE Clinical Senior Lectureship to AWM, a Versus Arthritis Foundation Fellowship (grant code 19764), the Wellcome Trust Institutional Strategic Support Fund to JIR and MYMY (204825/Z/16/Z) and NIHR Doctoral Research Fellowship to MYMY (DRF-2014-07-155).

## Author Contribution

Conceptualisation: JIR, MYMY, EMV, AWM; Methodology: JIR, MYMY, JHB, EMV, AWM; Data Collection and Resources: DLM, LL, ACR, MHB, DP, HJC, JDI, INB, PE, AB, TJV; Performing Experiments: JIR, MYMY, DW, MM, YES; Data Analysis: JIR, MYMY, VD, DW, MM, JHB, JT, AWM; Writing Original Draft: JIR, MYMY, EMV, AWM. All authors had full access to the data and approved the manuscript prior to submission to the journal.

## Competing Interest

Dr Vital and Prof Emery have received honoraria and consulting fees from Roche within the last 3 years. All other authors declare no competing interest related to the work described in this manuscript.

## Supplementary File

### Supplementary Materials and Methods

#### Study Population: Rheumatoid arthritis

251/611 patients with RA were recruited from the British Society for Rheumatology Biologics Register for RA (BSRBR-RA), as previously described (*50*). In this registry, collaborations were also established between large referring centres to form the Biologics in RA Genetics and Genomics Study Syndicate (BRAGGSS), which collected blood samples from bDMARD-treated RA patients (www.braggss.co.uk). A further 360 RA patients were recruited from the LTHT biologics clinic, as previously described (*40*). Inclusion criteria were a consultant diagnosis of RA (*51*); adults ((>18 years) at symptom onset of RA; and at least 6 months follow-up after rituximab therapy with documentation of clinical response. For functional studies, peripheral blood were obtained from three separate cohorts from LTHT; (i) DMARD-experienced adult RA patients with long-standing disease (>2 years), as previously published (*52*), ii) DMARD-naïve early RA patients (i.e. symptom duration <1 year) who were recruited into the Infliximab as Induction Therapy in Early RA (IDEA) study (*53*); and (iii) rituximab-treated RA patients from biologics clinic, where blood was taken immediately prior to first rituximab cycle.

#### Clinical outcomes: Rheumatoid arthritis

Clinical outcome measurements (SJC28, TJC28 and CRP, in mg/L) were taken at baseline (up to 6-weeks before the individual’s rituximab treatment) and again at follow-up (6 months from the individual’s first rituximab infusion, or 3 months if either the six month data were not available or the patient had received a further rituximab infusion prior to 6 months). Where CRP levels were recorded as below 5 mg/L, the lower limit of reliable detection, these were replaced by imputed values from a Uniform distribution from 0 to 5. Disease activity scores were calculated using the formula: 1.10 x [(0.56 x √TJC28) + (0.28 x SJC28) + (0.36 ln(CRP+1))] for the 3C-DAS28CRP (*54*) and [√SJC28 + (0.6 x ln (CRP + 1))] for the recently reported 2C-DAS28 (*35*).

#### Study population: Systemic lupus erythematosus

177/262 patients with SLE were recruited from the BILAG Biologics Registry (BILAG-BR) (*8*). While the other 85 patients were recruited from the connective tissue disease clinic, LTHT (*55*). Inclusion criteria were adults (≥18 years); fulfilling the revised 1997 American College of Rheumatology classification for SLE (*56*); active SLE as defined by 1 x BILAG A or 2 x BILAG B grades; and at least 6 months follow-up post-rituximab with documentation of clinical response. For the functional studies, peripheral blood was obtained immediately prior to first cycle rituximab in some SLE patients, as detailed above.

#### Clinical outcomes: Systemic lupus erythematosus

Disease activity was assessed using the BILAG-2004 index (*57*) at baseline and ∼ every 6 months thereafter. Clinical responses at 6 months were standardised between the two cohorts and determined as follows: (i) MCR = improvement of all domains rated A/B to grade C/better and no A/B flare between baseline and 6 months; (2) PCR = maximum of 1 domain with a persistent grade B with improvement in all other domains and no A or B flare; and (3) non-response = those not meeting the criteria for either MCR or PCR (*55, 58*). Global BILAG score was calculated as follows: grade A=12, grade B=8, grade C=1 and grades D and E=0 (*59*).

#### Treatment

All patients received a first cycle of therapy consisting of 100 mg of methylprednisolone and 1000 mg of rituximab given intravenously on days 1 and 14, with the exception of 3 RA patients who received 1g in total. All RA patients received rituximab MabThera® while 7/262 SLE patients received a biosimilar. Continuation of a stable dose or reduction of concomitant DMARDs and/or oral prednisolone was left to the clinicians’ discretion with the aim to stop glucocorticoids if remission was achieved within 6 months.

#### Routine Laboratory assessments

All autoantibody and immunoglobulin assessments were determined using standard assays in the routine NHS diagnostics laboratory of each participating site. RA patients who ever had RF and ACPA titres of >40 iu/mL and >7 iu/mL, respectively, were defined as positive, to maintain consistency with previously published studies. For SLE, ANA was tested using indirect immunofluorescence and a panel of nuclear autoantibodies including anti-dsDNA and anti-ENA antibodies (Ro52, Ro60, La, Sm, and RNP). Complement (C3 and C4) and total serum immunoglobulin (IgM, IgA and IgG) titres were measured by nephelometry. Adult reference ranges are as follows: C3: 0.75–1.65 g/L, C4: 0.14–0.54 g/L), IgG (6-16 g/L), IgA (0.8-4 g/L) and IgM (0.5-2 g/L).

#### FCGR2C copy number assay

High sequence identity between *FCGR2A*, *FCGR2B* and *FCGR2C* prevents the MLPA probes in panels P110 and P111 from uniquely recognising the copy number variable *FCGR2C* without simultaneously hybridising to *FCGR2B* or *FCGR2A*. The resulting interpretation of *FCGR2C* gene copy number requires combinations of multiple probe intensities.

Using a gene specific resequencing approach in 32 individual genomes we surveyed all *FCGR2* variants and classified them as either PSVs or SNPs. Long range PCR was used to amplify each gene specifically using the oligonucleotide primers in Supplementary Table 4. We identified a single PSV that differentiated all three genes (Supplementary Figure 2A). To supplement the MLPA panels we developed a novel assay to measure the copy number of *FCGR2C* directly referenced to both *FCGR2A* and *FCGR2B*. A single pair of primers was used to co-amplify 279bp fragments of all three genes with equal efficiency in a single reaction. PCR cycles (annealing at 55°C, 1.5mM MgCl2) were limited to 30, to ensure termination in the linear phase and to maintain proportional amplification of all three genes. Amplicons were purified and concentrated and normalised using a modified ChargeSwitch process, utilising a limiting quantity of magnetic beads and eluting in a fifth of the original volume. Purified amplicons were Sanger sequenced using the reverse primer, and relative electropherogram peak heights of gene specific PSVs were determined using QSVanalyser software (*60*). Comparison of the 2C-derived peak height with the invariant 2B and 2A, allowed copy number estimation with respect to two reference genes (Supplementary Figure 2B). Copy number of *FCGR2C* was inferred from cluster centres as indicated.

DNA extraction methods varied between the cohorts which led to some MLPA probes performing less well than others.

---

**Supplementary Figure 1.**
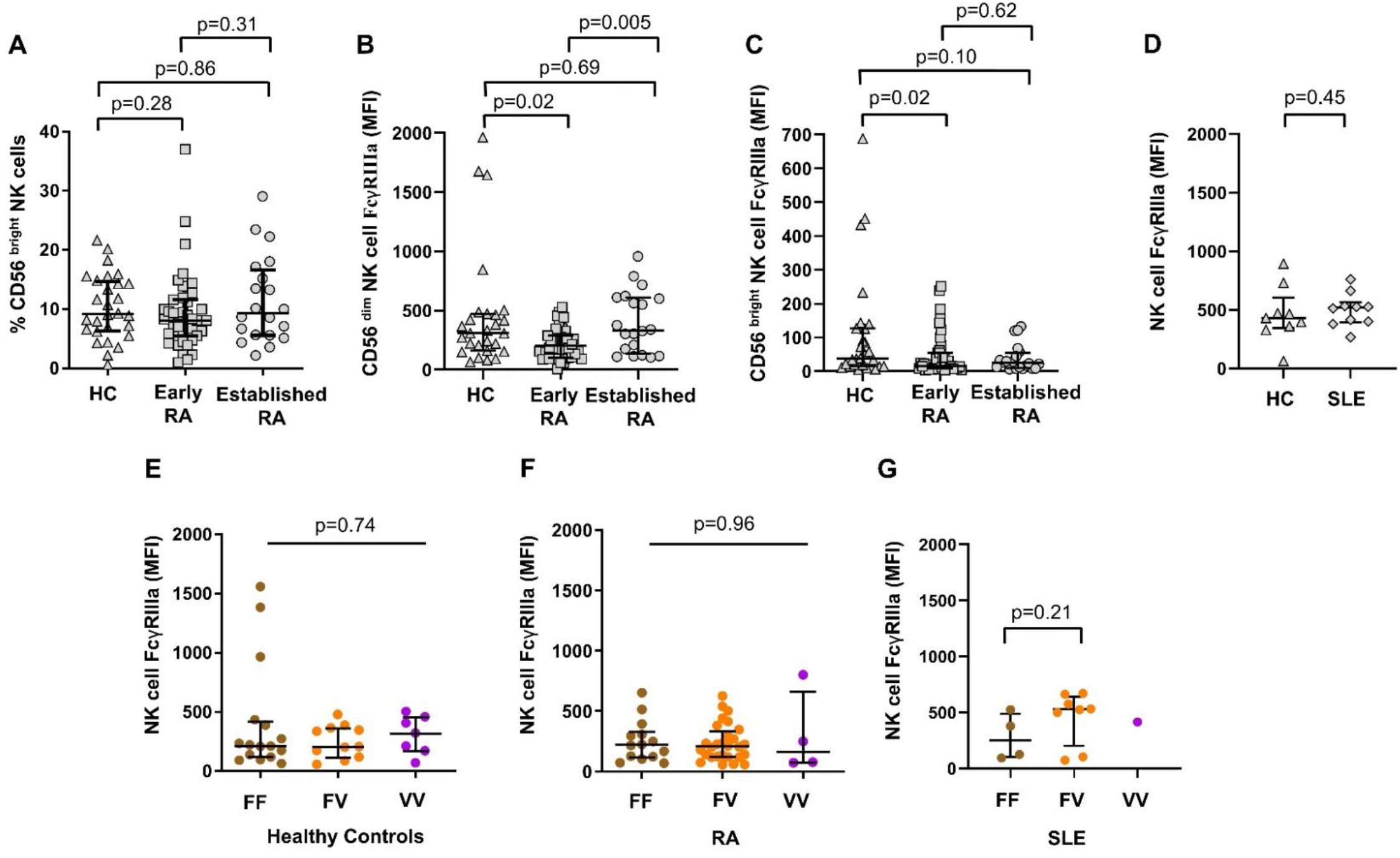
FcγRIIIa expression on natural killer cell subsets in healthy controls, rheumatoid arthritis and systemic lupus erythematosus. **(A)** Natural killer (NK) cell (CD3-CD56+CD16+) FcγRIIIa (CD16 clone 3G8) geometric mean fluorescence intensity (MFI) using flow cytometry for healthy controls (HC), early (symptom onset <1 year and treatment naïve) and established (>2 years) rheumatoid arthritis (RA). **(B)** Percentage of NK cells in the CD56^bright^ subset for HC, early and established RA. **(C)** FcγRIIIa expression on CD56^dim^ NK cells for HC, early and established RA. **(D)** FcγRIIIa expression on CD56^bright^ NK cells for HC, early and established RA. **(E)** NK cell FcγRIIIa expression in HC and SLE patients prior to rituximab treatment. FcγRIIIa expression on NK cells of different *FCGR3A*-F158V genotypes in HC **(F)**, RA **(G)** and SLE patients **(H)**, with two copies of *FCGR3A*. P-values calculated using non-parametric Mann-Whitney (bars with dropdowns) and Kruskal-Wallis H test (bars without dropdowns). Data are summarized using median and interquartile range.

**Supplementary Figure 2.**
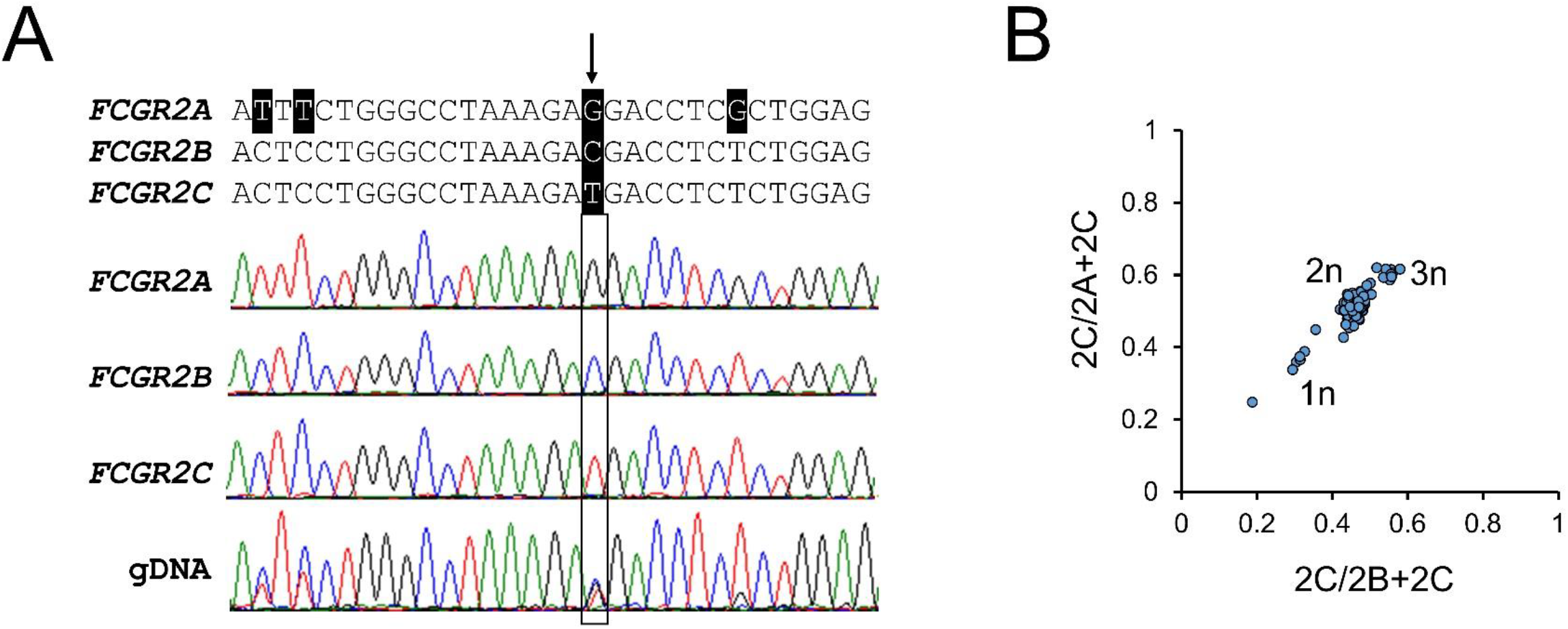
Novel *FCGR2C* QSV copy number assay. *FCGR2C QSV* exploits a unique paralogous sequence variant (arrowed) discriminating *FCGR2A*, *B* and *C* (hg19 Chr1:161,481,793; 161,645,283: 161,563,452 respectively). This variant is also known as NCBI dbSNP rs569867770. **(A)** Simultaneous PCR amplification of all three genes using common primers enabled proportional quantification of copy number variable *FCGR2C* with reference to the copy number invariable *FCGR2A* and *FCGR2B* from Sanger sequencing electropherograms using QSVanalyser. **(B)** Copy number calls based on the observed clustering were used to interpret *FCGR2C* STP/ORF quantitative genotypes from multiplex ligand proximity assays.

**Supplementary Figure 3.**
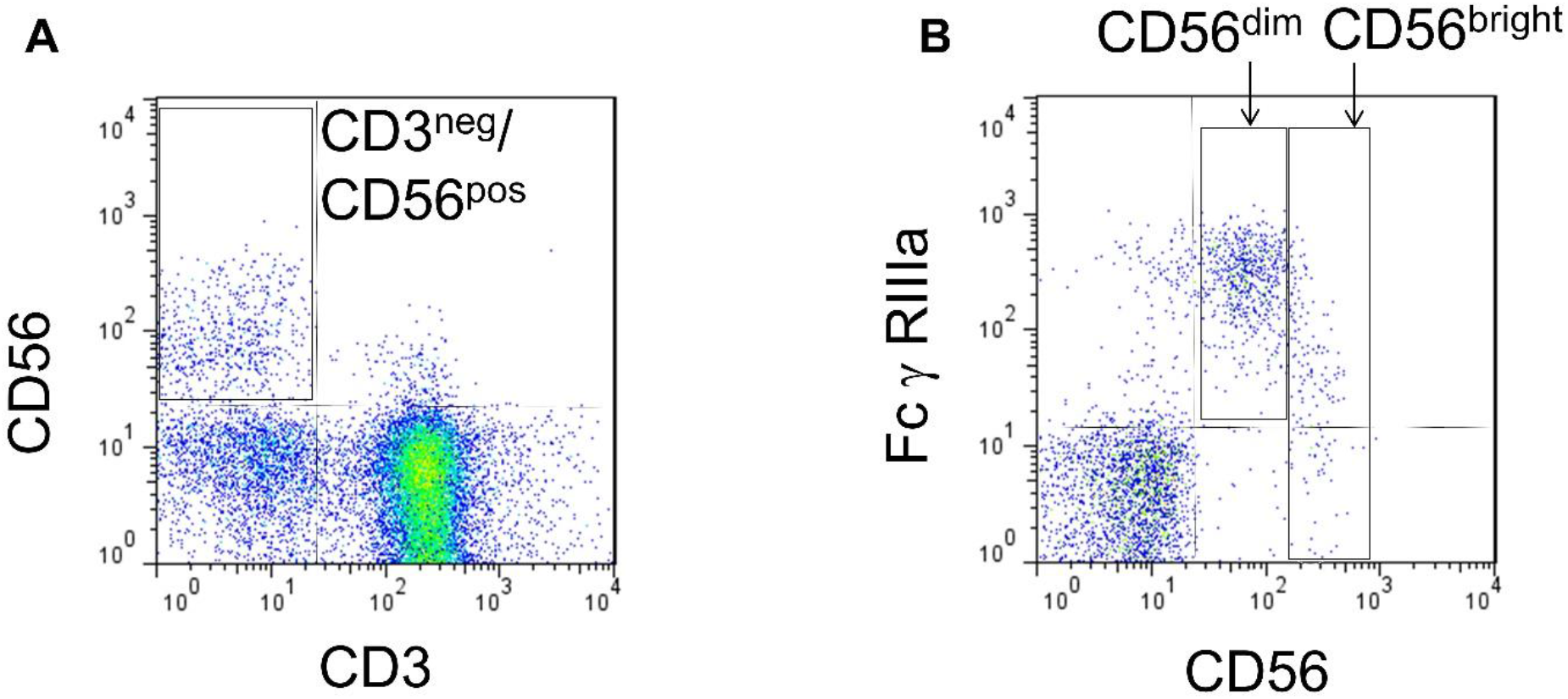
Natural Killer cell flow cytometric analysis. **(A)** Peripheral blood natural killer (NK) cells were identified on the basis of lymphocyte FSC/SSC characteristics and gating for lack of CD3 expression and positive expression of CD56 (CD3^neg^/CD56^pos^). **(B)** NK cell subsets were distinguished by separate gates created around CD56^dim^/CD16^++^ and CD56^bright^/CD16^neg/low^ NK cells.

**Supplementary Figure 4.**
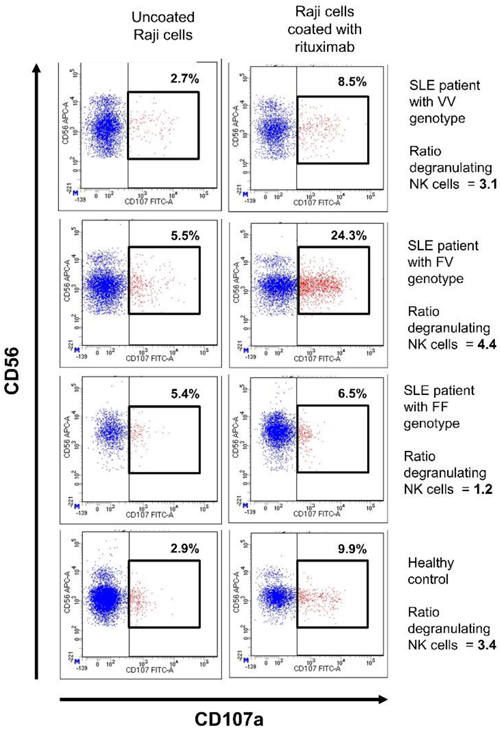
Natural Killer cell-mediated antibody dependent cellular cytotoxicity assay. The figure shows cell surface CD107a, gated on CD56+CD3-natural killer (NK) cells within peripheral blood mononuclear cells when co-cultured with Raji target cells using E:T ratio of 1:1 and in the absence of rituximab (left hand panel) and the presence of rituximab (right hand panel). The percentage values indicate the proportion of CD107+NK cells for each *FCGR3A* genotype in systemic lupus erythematosus (SLE) and healthy controls. Degranulation activity was measured by the ratio of NK cell degranulation with and without rituximab.

**Supplementary Table 1:**
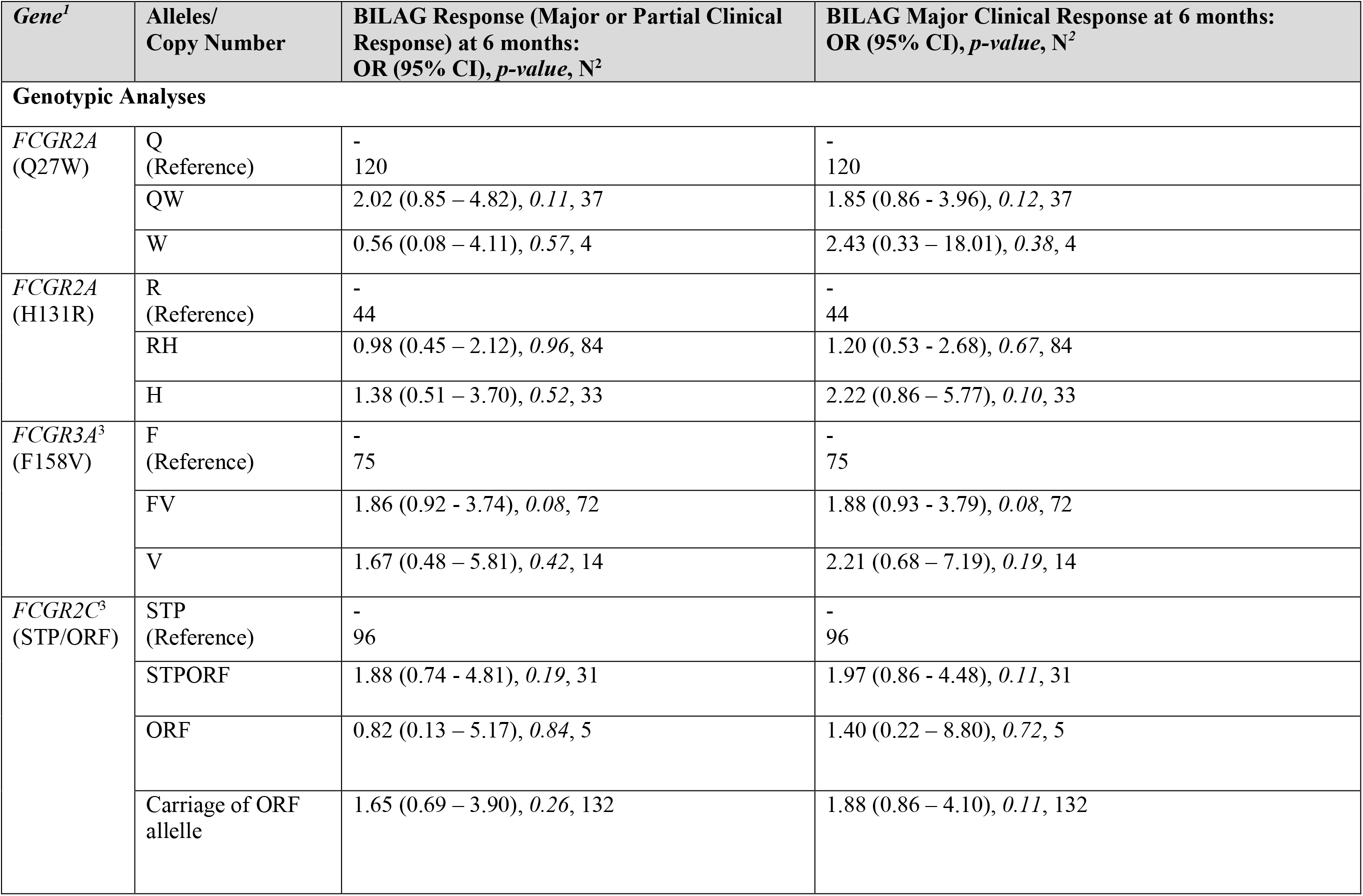
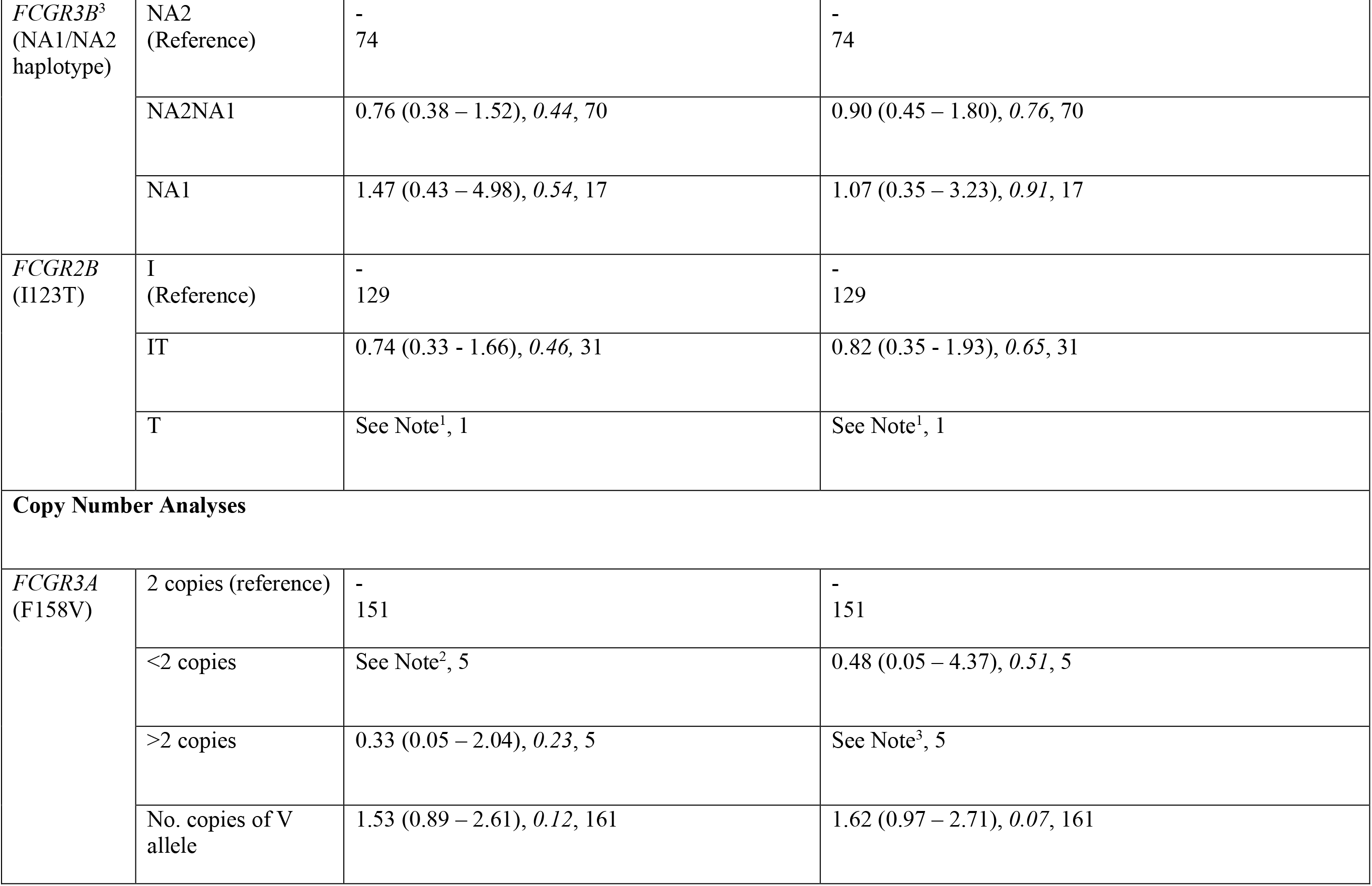
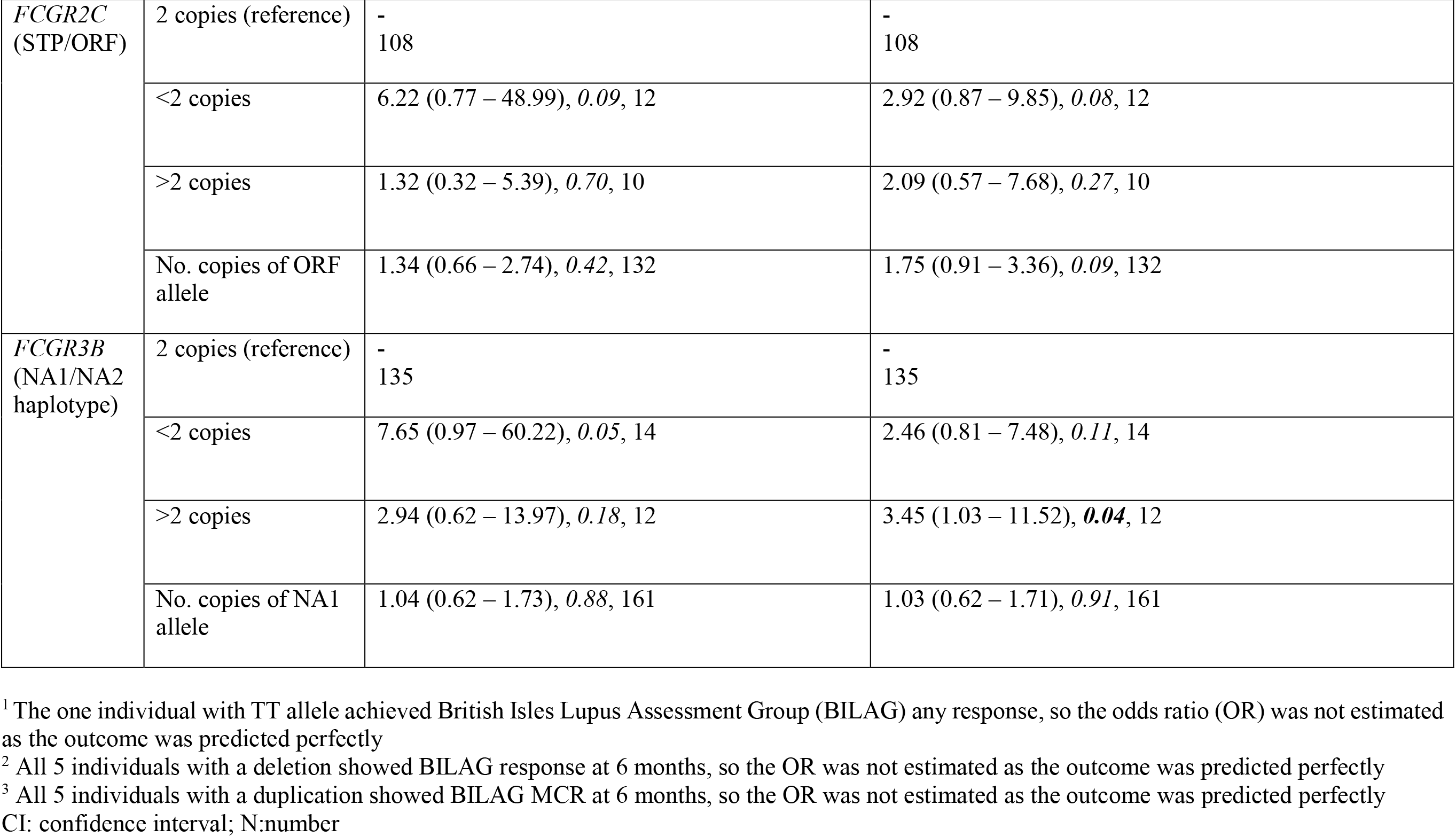
Effect of *FCGR* genotype and copy number on clinical response to rituximab in Caucasians with systemic lupus erythematosus

| *Gene*<sup>1</sup> | Alleles/<br>Copy Number | BILAG Response (Major or Partial Clinical Response) at 6 months:<br>OR (95% CI), *p*-value, N<sup>2</sup> | BILAG Major Clinical Response at 6 months:<br>OR (95% CI), *p*-value, N<sup>2</sup> |
| --- | --- | --- | --- |
| **Genotypic Analyses** |  |  |  |
| *FCGR2A*<br>(Q27W) | Q<br>(Reference) | -<br>120 | -<br>120 |
|  | QW | 2.02 (0.85 – 4.82), 0.11, 37 | 1.85 (0.86 - 3.96), 0.12, 37 |
|  | W | 0.56 (0.08 – 4.11), 0.57, 4 | 2.43 (0.33 – 18.01), 0.38, 4 |
| *FCGR2A*<br>(H131R) | R<br>(Reference) | -<br>44 | -<br>44 |
|  | RH | 0.98 (0.45 – 2.12), 0.96, 84 | 1.20 (0.53 - 2.68), 0.67, 84 |
|  | H | 1.38 (0.51 – 3.70), 0.52, 33 | 2.22 (0.86 – 5.77), 0.10, 33 |
| *FCGR3A*<sup>3</sup><br>(F158V) | F<br>(Reference) | -<br>75 | -<br>75 |
|  | FV | 1.86 (0.92 - 3.74), 0.08, 72 | 1.88 (0.93 - 3.79), 0.08, 72 |
|  | V | 1.67 (0.48 – 5.81), 0.42, 14 | 2.21 (0.68 – 7.19), 0.19, 14 |
| *FCGR2C*<sup>3</sup><br>(STP/ORF) | STP<br>(Reference) | -<br>96 | -<br>96 |
|  | STPORF | 1.88 (0.74 - 4.81), 0.19, 31 | 1.97 (0.86 - 4.48), 0.11, 31 |
|  | ORF | 0.82 (0.13 – 5.17), 0.84, 5 | 1.40 (0.22 – 8.80), 0.72, 5 |
|  | Carriage of ORF<br>allele | 1.65 (0.69 – 3.90), 0.26, 132 | 1.88 (0.86 – 4.10), 0.11, 132 |

| *Gene*<sup>1</sup> | Alleles/<br>Copy Number | BILAG Response (Major or Partial Clinical<br>Response) at 6 months:<br>OR (95% CI), *p*-value, N<sup>2</sup> | BILAG Major Clinical Response at 6 months:<br>OR (95% CI), *p*-value, N<sup>2</sup> |
| --- | --- | --- | --- |
| *FCGR3B*<sup>3</sup><br>(NA1/NA2<br>haplotype) | NA2<br>(Reference) | -<br>74 | -<br>74 |
|  | NA2NA1 | 0.76 (0.38 – 1.52), 0.44, 70 | 0.90 (0.45 – 1.80), 0.76, 70 |
|  | NA1 | 1.47 (0.43 – 4.98), 0.54, 17 | 1.07 (0.35 – 3.23), 0.91, 17 |
| *FCGR2B*<br>(I123T) | I<br>(Reference) | -<br>129 | -<br>129 |
|  | IT | 0.74 (0.33 - 1.66), 0.46, 31 | 0.82 (0.35 - 1.93), 0.65, 31 |
|  | T | See Note<sup>1</sup>, 1 | See Note<sup>1</sup>, 1 |
| **Copy Number Analyses** |  |  |  |
| *FCGR3A*<br>(F158V) | 2 copies (reference) | -<br>151 | -<br>151 |
|  | <2 copies | See Note<sup>2</sup>, 5 | 0.48 (0.05 – 4.37), 0.51, 5 |
|  | >2 copies | 0.33 (0.05 – 2.04), 0.23, 5 | See Note<sup>3</sup>, 5 |
|  | No. copies of V<br>allele | 1.53 (0.89 – 2.61), 0.12, 161 | 1.62 (0.97 – 2.71), 0.07, 161 |
| *FCGR2C*<br>(STP/ORF) | 2 copies (reference) | -<br>108 | -<br>108 |
|  | <2 copies | 6.22 (0.77 – 48.99), 0.09, 12 | 2.92 (0.87 – 9.85), 0.08, 12 |
|  | >2 copies | 1.32 (0.32 – 5.39), 0.70, 10 | 2.09 (0.57 – 7.68), 0.27, 10 |
|  | No. copies of ORF allele | 1.34 (0.66 – 2.74), 0.42, 132 | 1.75 (0.91 – 3.36), 0.09, 132 |
| *FCGR3B*<br>(NA1/NA2 haplotype) | 2 copies (reference) | -<br>135 | -<br>135 |
|  | <2 copies | 7.65 (0.97 – 60.22), 0.05, 14 | 2.46 (0.81 – 7.48), 0.11, 14 |
|  | >2 copies | 2.94 (0.62 – 13.97), 0.18, 12 | 3.45 (1.03 – 11.52), **0.04**, 12 |
|  | No. copies of NA1 allele | 1.04 (0.62 – 1.73), 0.88, 161 | 1.03 (0.62 – 1.71), 0.91, 161 |

<sup>1</sup> The one individual with TT allele achieved British Isles Lupus Assessment Group (BILAG) any response, so the odds ratio (OR) was not estimated as the outcome was predicted perfectly
<sup>2</sup> All 5 individuals with a deletion showed BILAG response at 6 months, so the OR was not estimated as the outcome was predicted perfectly
<sup>3</sup> All 5 individuals with a duplication showed BILAG MCR at 6 months, so the OR was not estimated as the outcome was predicted perfectly CI: confidence interval; N:number

**Supplementary Table 2:** Baseline clinical characteristics and laboratory measures and association with complete B cell depletion in the combined rheumatoid arthritis and systemic lupus erythematosus analyses

| Characteristics | Sample Size | Complete B cell depletion, Mean (SD) or number (%) positive | Incomplete B cell depletion, Mean (SD) or number (%) positive | Complete B cell depletion post-RTX, OR (95% CI), *p-value* |
| --- | --- | --- | --- | --- |
| Age at first RTX cycle (years, effect per 10 years) | 394 | 5.61 (1.44) | 5.26 (1.57) | 1.17 (1.02 – 1.34), **0.02** |
| Sex (Female) | 413 | 206 (87%) | 141 (80%) | 1.75 (1.03 – 2.98), **0.04** |
| Disease indication [i.e. RA as Reference] | 387 | 44 (52%) | 41 (48%) | 0.76 (0.47 – 1.23), 0.26 |
| Concomitant DMARDs<sup>1</sup>, including HCQ | 387 | 189 (86%) | 123 (74%) | 2.18 (1.31-3.64), **0.003** |
| Concomitant oral prednisolone<sup>2</sup> | 85 | 33 (75%) | 30 (73.2%) | 1.10 (0.42 – 2.90), 0.85 |
| Oral prednisolone dose (mg/day)<sup>2</sup> | 85 | 13.7 (12.6) | 13.6 (11.6) | 1.00 (0.97 – 1.04), 0.96 |
| IgM (g/L) | 397 | 1.4 (0.7) | 1.5 (1.1) | 0.84 (0.67 – 1.05), 0.13 |
| IgA (g/L) | 398 | 3.0 (1.3) | 3.4 (1.6) | 0.82 (0.71 – 0.95), **0.01** |
| IgG (g/L) | 398 | 12.1 (4.1) | 13.6 (5.2) | 0.93 (0.89 – 0.98), **0.003** |
| Total B cell counts (x 10<sup>9</sup>/L)<sup>3</sup> | 395 | 124.3 (125) | 143.4 (145) | 1.00 (1.00 – 1.00), 0.16 |
| **Naïve B cell counts (x 10<sup>9</sup>/L)<sup>3</sup>** | 393 | 93.3 (96) | 106.2 (123) | 1.00 (1.00 – 1.01), *0.25* |
| **Memory B cell counts (x 10<sup>9</sup>/L)<sup>3</sup>** | 393 | 28.6 (54) | 32.1 (34) | 1.00 (0.99 – 1.00), *0.47* |
| **Plasmablast counts (x 10<sup>9</sup>/L)<sup>3</sup>** | 395 | 2.3 (3) | 5.8 (9) | 0.84 (0.78 – 0.89), *<0.001* |

<sup>1</sup> Concomitant disease modifying anti-rheumatic drugs (DMARDs)
<sup>2</sup> Data available for systemic lupus erythematosus (SLE) Leeds cohort only
<sup>3</sup> Count x 10<sup>9</sup> cells/L for each subset multiplied by 1000 prior to analysis

**Supplementary Table 3.** Effect of *FCGR* genotype and copy number on early complete B cell depletion following rituximab in rheumatoid arthritis and systemic lupus erythematosus

| *Gene*<sup>1</sup> | Alleles/<br>Copy Number | Complete B cell<br>Depletion:<br>N (%) | Incomplete B cell<br>Depletion:<br>N (%) | Unadjusted Complete B cell<br>Depletion post-RTX:<br>OR (95% CI), *p-value*<sup>2</sup> | Adjusted Complete B cell<br>Depletion post-RTX<sup>3</sup>:<br>OR (95% CI), *p-value*<sup>2</sup> |
| --- | --- | --- | --- | --- | --- |
| **Genotypic Analyses** |  |  |  |  |  |
| *FCGR2A*<br>(Q27W) | Q<br>(Reference) | 173 (73.3) | 138 (78.0) | - | - |
|  | QW | 59 (25.0) | 34 (19.2) | 1.38 (0.86 – 2.23), 0.18 | 1.22 (0.71 – 2.11), 0.46 |
|  | W | 4 (1.7) | 5 (2.8) | 0.64 (0.17 – 2.42), 0.51 | 1.28 (0.27 – 5.99), 0.76 |
| *FCGR2A*<br>(H131R) | R<br>(Reference) | 63 (26.7) | 47 (26.6) | - | - |
|  | RH | 114 (48.3) | 94 (53.1) | 0.90 (0.57 – 1.44), 0.67 | 0.74 (0.43 – 1.27), 0.27 |
|  | H | 59 (25.0) | 36 (20.3) | 1.22 (0.70 – 2.14), 0.48 | 1.29 (0.67 – 2.48), 0.44 |
| *FCGR3A*<sup>4</sup><br>(F158V) | F<br>(Reference) | 95 (40.3) | 86 (48.6) | - | - |
|  | FV | 114 (48.3) | 80 (45.2) | 1.29 (0.86 – 1.94), 0.22 | 1.13 (0.70 – 1.82), 0.61 |
|  | V | 27 (11.4) | 11 (6.2) | 2.22 (1.04 – 4.75), **0.04** | 3.03 (1.23 – 7.42), **0.02** |
| *FCGR2C*<sup>4</sup><br>(STP/ORF) | STP<br>(Reference) | 156 (70.0) | 126 (75.0) | - | - |
|  | STPORF | 59 (26.5) | 36 (21.4) | 1.32 (0.82 – 2.13), 0.25 | 1.28 (0.75 – 2.21), 0.36 |
|  | ORF | 8 (3.6) | 6 (3.6) | 1.08 (0.36 – 3.18), 0.89 | 1.51 (0.41 – 5.58), 0.54 |
| *FCGR3B*<sup>4</sup><br>(NA1/NA2<br>haplotype) | NA2<br>(Reference) | 98 (44.0) | 65 (40.4) | - | - |
|  | NA2NA1 | 98 (44.0) | 75 (46.6) | 0.87 (0.56 – 1.34), 0.52 | 1.04 (0.62 - 1.74), 0.88 |
|  | NA1 | 27 (12.1) | 21 (13.0) | 0.85 (0.44 – 1.63), 0.63 | 0.74 (0.36 – 1.55), 0.43 |
| *FCGR2B*<br>(I123T) | I<br>(Reference) | 184 (78.3) | 137 (78.3) | - | - |
|  | IT | 48 (20.4) | 36 (20.6) | 0.99 (0.61 – 1.61), 0.98 | 0.87 (0.49 - 1.54), 0.63 |
|  | T | 3 (1.3) | 2 (1.1) | 1.12 (0.18 – 6.78), 0.90 | 0.43 (0.06 – 3.21), 0.41 |
| **Copy Number Analyses** |  |  |  |  |  |
| *FCGR3A*<br>(F158V) | 2 copies<br>(reference) | 208 (88.1) | 165 (93.2) | - | - |
|  | <2 copies | 5 (2.1) | 1 (0.6) | 3.97 (0.46 – 34.28), 0.21 | 3.12 (0.33 – 29.57), 0.32 |
|  | >2 copies | 23 (9.8) | 11 (6.2) | 1.66 (0.79 – 3.50), 0.18 | 2.35 (0.99 – 5.57), **0.05** |
|  | No. copies of V<br>allele | 141 (59.7) | 91 (51.4) | 1.41 (1.05 – 1.90), **0.02** | 1.49 (1.06 – 2.08), **0.02** |
| *FCGR2C*<br>(STP/ORF) | 2 copies<br>(reference) | 172 (77.1) | 135 (80.4) | - | - |
|  | <2 copies | 22 (9.9) | 12 (7.1) | 1.44 (0.69 – 3.01), 0.33 | 1.23 (0.52 – 2.88), 0.64 |
|  | >2 copies | 29 (13.0) | 21 (12.5) | 1.08 (0.59 – 1.99), 0.79 | 1.28 (0.65 – 2.54), 0.48 |
|  | No. copies of<br>ORF allele | 67 (30.0) | 42 (25.0) | 1.23 (0.87 – 1.75), 0.24 | 1.35 (0.90 – 2.02), 0.15 |
| *FCGR3B*<br>(NA1/NA2<br>haplotype) | 2 copies<br>(reference) | 191 (80.9) | 146 (82.5) | - | - |
|  | <2 copies | 22 (9.3) | 14 (7.9) | 1.20 (0.59 – 2.43), 0.61 | 1.13 (0.51 – 2.54), 0.76 |
|  | >2 copies | 23 (9.8) | 17 (9.6) | 1.03 (0.53 – 2.01), 0.92 | 1.14 (0.54 – 2.40), 0.73 |
|  | No. copies of<br>NA1 allele | 131 (57.2) | 104 (61.5) | 0.85 (0.64 – 1.14), 0.28 | 0.91 (0.65 – 1.27), 0.56 |

<sup>1</sup>Genes presented in chromosomal order on 1q23 centromere to telomere.
<sup>2</sup>Odds ratio (OR), 95% confidence intervals (CI) and p-value for the effect of the indicated genotype or copy number on complete B-cell depletion at 2 weeks, compared with reference genotype. All tests were performed using univariable logistic regression.
<sup>3</sup>Analyses adjusted for age, concomitant disease-modifying anti-rheumatic drug, including hydroxychloroquine, and baseline plasmablast count.
<sup>4</sup>*FCGR3A*, *FCGR2C* and *FCGR3B* are subject to copy number variation, analyses were performed according to biallelic genotype whereby the effect of heterozygosity and homozygosity for the rare allele were compared with homozygosity for the common allele.
N: number

**Supplementary Table 4.**
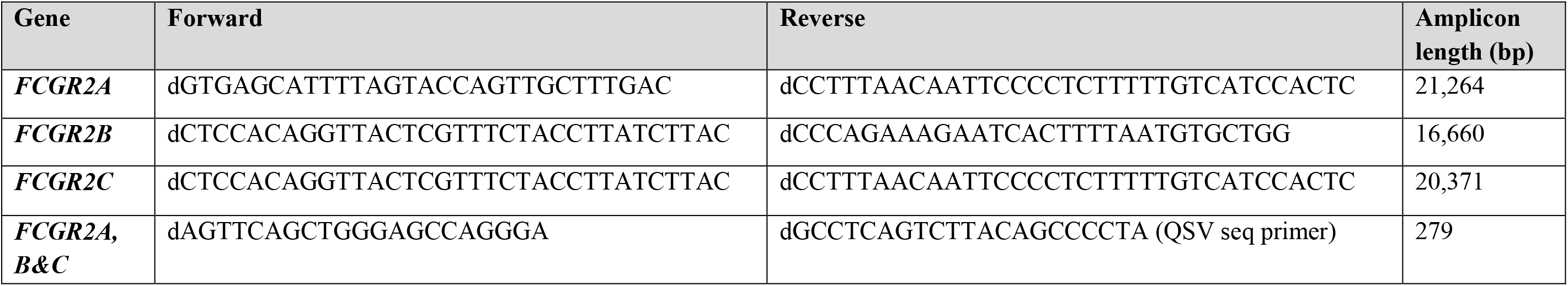
Oligonucleotide primer sequences used to resequence *FCGR2* genes and a novel *FCGR2C* QSV assay

| Gene | Forward | Reverse | Amplicon length (bp) |
| --- | --- | --- | --- |
| *FCGR2A* | dGTGAGCATTTTAGTACCAGTTGCTTTGAC | dCCTTTAACAATTCCCCTCTTTTTGTCATCCACTC | 21,264 |
| *FCGR2B* | dCTCCACAGGTTACTCGTTTCTACCTTATCTTAC | dCCCAGAAAGAATCACTTTTAATGTGCTGG | 16,660 |
| *FCGR2C* | dCTCCACAGGTTACTCGTTTCTACCTTATCTTAC | dCCTTTAACAATTCCCCTCTTTTTGTCATCCACTC | 20,371 |
| *FCGR2A, B&C* | dAGTTCAGCTGGGAGCCAGGGA | dGCCTCAGTCTTACAGCCCCTA (QSV seq primer) | 279 |

## Appendix I MATURA Consortium

**Work Stream 1:** Prof Costantino Pitzalis (Queen Mary University of London), Prof Peter Taylor (University of Oxford), Prof Ernest Choy (Cardiff University), Prof Iain McInnes (University of Glasgow), Dr Mike Barnes (Queen Mary University of London), Prof John Isaacs (Newcastle University), Prof Christopher Buckley (University of Birmingham), Prof Michael Ehrenstein (University College London), Prof Peter Sasieni (Queen Mary University of London), Dr Andrew Filer (University of Birmingham).

**Work Stream 2:** Prof Anne Barton (University of Manchester), Prof Ann Morgan (University of Leeds), Prof Gerry Wilson (University College Dublin), Prof Paul McKeigue (University of Edinburgh), Prof Heather Cordell (Newcastle University), Prof Jenny Barrett (University of Leeds), Prof Andrew Cope (Kings College London), Prof Adam Young (University of Hertfordshire), Prof Karim Raza (University of Birmingham), Prof Katherine Payne (University of Manchester), Prof Jane Worthington (University of Manchester), Prof Deborah Symmons (University of Manchester), Prof Kimme Hyrich (University of Manchester). Industry: Martin Hodge (Pfizer), Anthony Rowe (Janssen), Jianmei Wang (Roche/Genentech), Michelle Mao (BGI), Patricia McLoughlin (Qiagen), Carolyn Cuff (AbbVie), David Close (MedImmune).

**Leeds Biologics Service (University of Leeds and Leeds Teaching Hospitals NHS Trust)**

**Management Team:** Paul Emery, Maya Buch, Elizabeth MA Hensor

**Consultant and Senior Scientific Staff:** Maya Buch, Paul Emery, Ann Morgan, Frederique Ponchel, Shouvik Dass, Edward Vital, Sarah Bingham

**Specialist Registrars and Clinical Fellows:** Edith Villeneuve, Sudipto Das, Jacqueline Nam, Sarah Horton, Sarah Mackie, Rebecca Thomas, Lesley-Anne Bissell, Chadi Rakieh, Zoe Ash, Sarah Twigg, Laura Coates, Fahad Fazal, Laura Hunt, Esme Ferguson, Sara Else, Gui Tran, Ahmed Zayat, Guiseppina Abignano, Md Yuzaiful Md Yusof, Radhika Raghunath, Hannah Mathieson, Chitra Salem-Ramakumaran, Hanna Gul, Mahwish Mahmood, Leticia Montoya Garcia, Jean Baptiste Candelier, Thibault Rabin, Gisela Eugenio, Joana Fonseca Ferreira.

**Biologics Nursing Staff:** Pauline Fitzgerald, Matthew Robinson, Jason Ward, Beverly Wells, David Pickles, Oliver Wordsworth, Christine Thomas, Alison McManus, Lynda Bailey, Linda Gray, Kate Russell, Jayne Davies

**Laboratory and Support Staff:** Diane Corscadden, Karen Henshaw, Katie Mbara, Stephen Martin, James Robinson, Dawn Wild, Agata Burska, Sarah Fahey, Jill Halsted-Rastrick, Ged Connoly-Thompson, Jonathan Thompson, Ian Weatherill, Andrea Paterson

**IACON Radiology team:** Laura Horton, Alwyn Jackson, Richard Hodgson

## Appendix II Contributors to the MASTERPLANS Consortium

The University of Manchester: Prof Katherine Payne; Dr Mark Lunt; Prof Niels Peek; Dr Nophar Geifman; Dr Sean Gavan; Dr Gillian Armitt; Dr Patrick Doherty; Dr Jennifer Prattley; Dr Narges Azadbakht; Angela Papazian; Dr Helen Le Sueur; Carmen Farrelly; Clare Richardson; Zunnaira Shabbir; Lauren Hewitt; Dr Emily Sutton; Alison Fountain, Patrick Doherty.

University of Bath: Prof Neil McHugh.

University of Birmingham: Prof Caroline Gordon; Prof Stephen Young.

University of Cambridge: Prof David Jayne; Prof Vern Farewell; Dr Li Su.

Imperial College London: Prof Matthew Pickering; Prof Elizabeth Lightstone; Dr Alyssa Gilmore; Prof Marina Botto.

King’s College London: Prof Timothy Vyse; Dr David Lester Morris; Prof David D’Cruz.

University of Liverpool: Prof Michael Beresford; Prof Christian Hedrich; Dr Angela Midgley; Dr Jenna Gritzfeld.

University College London: Prof Michael Ehrenstein; Prof David Isenberg; Mariea Parvaz.

MASTERPLANS Patient and Public Involvement Group: Jane Dunnage; Jane Batchelor Elaine Holland; Pauline Upsal

